# Leveraging spatial structure to design spatially-targeted malaria interventions at the community-scale

**DOI:** 10.64898/2026.02.13.26346071

**Authors:** Michelle V Evans, Benjamin Roche, Vincent Herbreteau, Christophe Révillion, Thibault Catry, Matthew H. Bonds, Karen E. Finnegan, Ezra Mitsinjoniala, Felana A. Ihantamalala, Mauricianot Randriamihaja, Oméga Raobela, Andres Garchitorena

## Abstract

Progress in malaria control has stagnated since the early 21st century in many countries, requiring new approaches such as the use of spatially-targeted interventions. Evidence on the effectiveness of spatially-targeted interventions is mixed. Their success can be dependent on whether the setting is endemic, the metrics used to target the intervention, and the spatial resolution and scale of deployment. We developed a two-age-class, spatially-explicit model of malaria at the community-scale for a district in southeastern Madagascar, accounting for environmental heterogeneity and human mobility. The model was fit to field-based case notifications and malaria prevalence data and then used to simulate three interventions: indoor residual spraying (IRS), long-lasting insecticide-treated nets (LLIN), and active case detection (ACD). We compared five spatial targeting scenarios for each simulated intervention: (i) equally distributed, (ii) targeting communities nearest or (iii) furthest from clinics, (iv) targeting communities with highest incidence, and (v) targeting communities that are spatially central.

The non-targeted intervention was generally the most effective, but the least resource efficient. The second most effective intervention was based on spatial centrality, which reached a larger population while using fewer transportation resources than the equally distributed. No combination of interventions was able to eliminate malaria in the district, although a “perfect” ACD intervention could avert 100% of severe malaria cases. These results highlight the potential for targeted malaria interventions, especially in low-income settings, that take into account spatial structure in the human population and mobility to reduce malaria burden using fewer resources than conventional district-wide interventions.

## INTRODUCTION

Following nearly two decades of steady reductions in the global malaria burden, progress began to stall in the late 2010s and, in some countries, has since reversed (World Health Organization 2021, Oladipo et al. 2022). This has been due to a combination of factors. *Anopheles* mosquito vectors have become more resistant to the insecticide used in long-lasting insecticidal bednets (LLINs) and indoor residual spraying (IRS) campaigns (Ranson and Lissenden 2016), altering mosquito behavior to bite outside of homes, where exposure to insecticide is lessened (Sherrard-Smith et al. 2019). The *Plasmodium falciparum* parasite itself is becoming more resistant to treatments; first, to monotherapies such as chloroquine, and now to the arteminisinin-based combination therapy used globally to treat *P. falciparum* infections (Rosenthal et al. 2024). Increasing temperatures are shifting the endemic ranges of malaria, exposing new geographic regions and immune-naive populations to the disease, while reducing incidence rates in certain highly endemic regions (Carlson et al. 2023, Leal Filho et al. 2023). Global spending on malaria control has been consistently below the WHO-recommended target by nearly 50% (World Health Organization 2024), with funding expected to become even more limited given predicted reductions in development funding (Symons et al. 2025, Coughlan and Mehta 2025). New approaches are therefore needed to continue reducing malaria burdens globally and prevent losing recent progress while adapting to more limited resources.

One such approach is the use of spatially-targeted interventions, rather than “one-size-fits-all” national campaigns (Moonen et al. 2010, Greenwood 2017). However, there is conflicting evidence on the effectiveness of spatially-targeted malaria interventions (Khundi et al. 2021). Prior work has found that targeted interventions are most effective in elimination settings, where malaria persists in isolated patches (Pinchoff et al. 2015). The metric chosen to guide spatial-targeted intervention can also impact its effectiveness, with the proper metric dependent on the planned intervention (Cohen et al. 2017). In addition, the scale of the intervention and the geographic range of expected impact can influence spatially-targeted interventions measured impacts (Gosling et al. 2020, Lubinda et al. 2021, Benjamin-Chung et al. 2024). Specifically, down-scaling targeted interventions to allow district health teams to design and guide the interventions within their district has been proposed as one method to successfully implement spatially targeted campaigns while overcoming some of these weaknesses (Gosling et al. 2020).

Within a health district, community health workers (CHWs) play a particularly important role in providing care. As local members of the community, CHWs provide basic health services to small catchments of the population (Olaniran et al. 2017), and often serve an integral role in disease control programs (Win Han Oo et al. 2019). Due to their close proximity to patients, CHWs are well-positioned to conduct proactive care, such as active case detection (ACD) of malaria in patients with febrile illness (Stratil et al. 2021, Otambo et al. 2022), and encourage prophylaxis and preventive behaviors, such as intermittent preventive treatment during pregnancy (Enguita-Fernàndez et al. 2021, Koita et al. 2024). Finally, because they cover localized catchments, they are a natural implementer of spatially-targeted campaigns to reduce overall costs (Saint-Firmin et al. 2021), and have been used for spatially-explicit control of tuberculosis (Burke et al. 2021), maternal health program support (Gilmore and McAuliffe 2013), and pandemic response (Feroz et al. 2021). While there is potential for CHWs to implement community-scale, spatially-targeted malaria campaigns, this requires alignment among data and interventions at appropriately fine spatial scales to guide operations. Few studies have empirically modeled malaria transmission dynamics in response to interventions at this spatial scale across an entire health district to inform local decision-making.

Here, we develop a spatially-explicit dynamic model of malaria that includes spatial heterogeneity in human movement patterns and environmental forcing to evaluate the impact of different control interventions at the community-scale for a high transmission, rural district in southeastern Madagascar. The model includes fine resolution spatial data of human movement based on a field-derived mapping exercise of all footpaths in the district and climatic variables sourced from satellite imagery. We parameterize and fit the model to two independent datasets of malaria case rates and prevalence rates at the community-scale using Approximate Bayesian Computing (ABC). The fit model is used to compare five different scenarios of spatial targeting of three common interventions: long-lasting insecticidal nets (LLIN), indoor residual spraying (IRS), and active case detection (ACD). The resulting estimates of averted cases are integrated into an application that is available to local stakeholders that allows them to evaluate various intervention scenarios within the district.

## METHODS

### Study Setting

Ifanadiana district is a rural health district located in the Vatovavy region of southeastern Madagascar (Supp. Fig. 1). The district’s population of approximately 200,000 is divided into 15 communes and 195 fokontany (the smallest administrative unit comprising several villages) (INSTAT Madagascar 2021). One paved road runs east-west across the district, with an unpaved road connecting the northern and southern portions of the district. The majority of the population lives in rural villages that are inaccessible by vehicle and are located further than one hour from the nearest health center (Ihantamalala et al. 2020). Each commune contains at least one primary health center, and six communes contain an additional basic health center that provides more limited services, both of which are able to treat malaria cases of all-ages.

**Figure 1.**
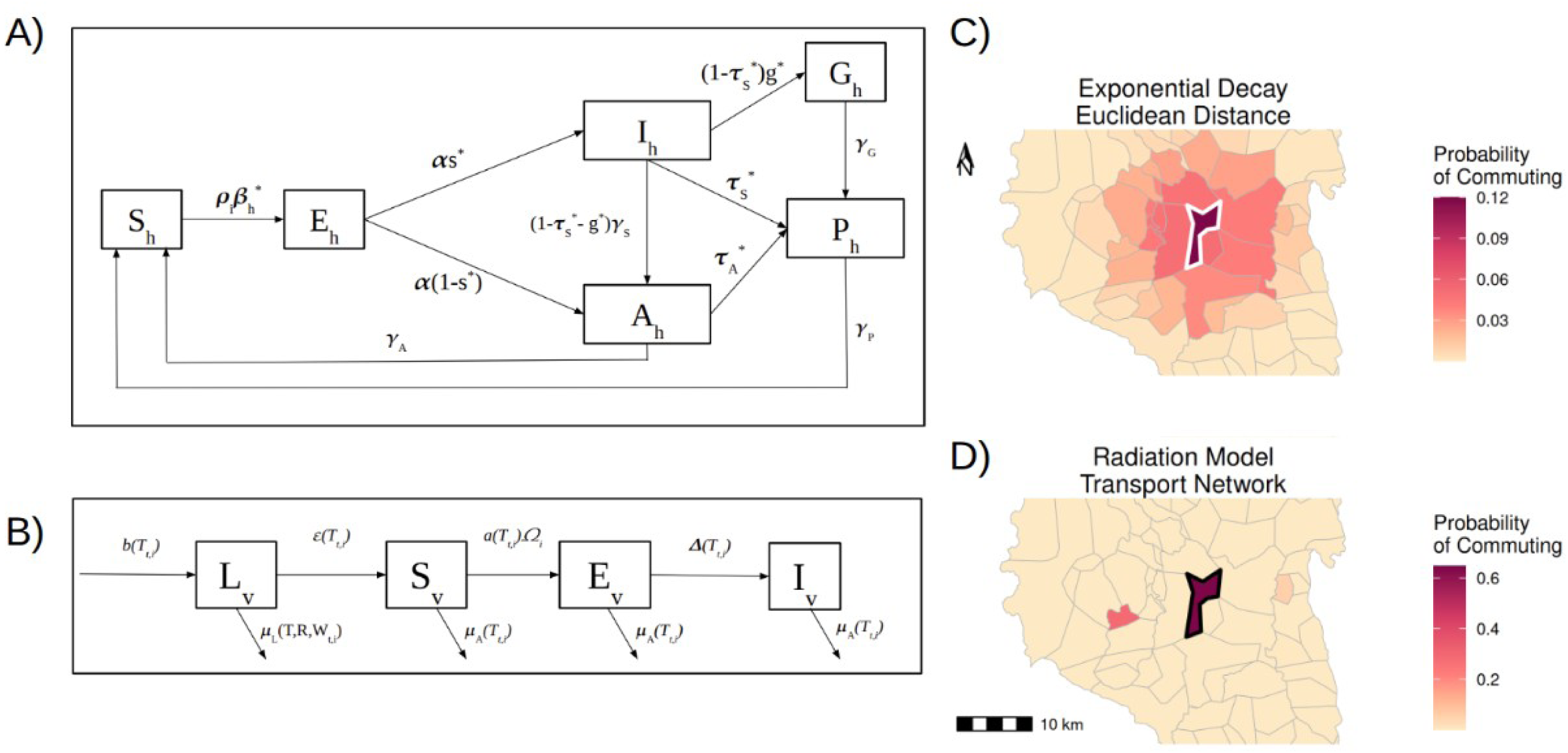
Diagram of compartmental model (A & B) and mobility models (C & D). A) Compartmental diagram of host compartments for one age-class. Parameters with a * superscript are age-specific. B) Compartmental diagram of vector compartments. Parameters represented as f() are a function of temperature T, rainfall R, or flooding W. C) Example metapopulation mobility model based on an exponential decay model on a euclidean distance network. Individuals are commuting from the patch (fokontany) highlighted in white. D) Example metapopulation mobility model based on a radiation model on a transport network from OpenStreetMap. Individuals are commuting from the patch (fokontany) highlighted in black. All parameters are defined in Table S1.

Each fokontany has a community health site staffed by community health workers who are able to treat malaria cases in children under 5 years old and pregnant women (Razafinjato et al. 2024). The district includes a strong elevational gradient from an altitude of 1400m in the west to 100m in the east. The landscape is characterized by a forest-savanna matrix interspersed with agricultural land used for rice production. Although the district only encompasses an area of 3970 km ^2^, this diversity of ecoregions results in spatio-temporal heterogeneities in malaria burdens across the district (Pourtois et al. 2023). A population-level survey conducted in 2021 found a district-level rate for children under 15 years old of 0.41, with village-level prevalence attaining rates as high as 0.8 (Evans et al. 2023).

Since 2015, Ifanadiana District has served as a model health district as part of a partnership between the Madagascar Ministry of Public Health and the non-governmental organization Pivot. This health system strengthening intervention includes improvements in system readiness and quality of care, a comprehensive monitoring and evaluation platform, and an actionable research program focused on outputs that can be used by local decision makers (Cordier et al. 2020). As part of the monitoring and evaluation platform, a district-representative, longitudinal cohort survey (Ifanadiana Health Outcomes and Prosperity longitudinal Evaluation, IHOPE) has been conducted every two years, collecting information on household demographics, disease incidence, and health system utilization (Miller et al. 2017). Data from the 2021 wave of this survey was used to inform treatment rates, symptomatic rates, baseline intervention levels, and malaria prevalence rates, described in detail below.

### Model Definition

We developed a dynamic model of host and vector dynamics that modeled patch-specific (e.g. fokontany-level) transition rates on a daily timestep from January 1 2006 - December 31 2021. Hosts were modeled via two age classes, children under-five and adults and children over five years old (referred to as “adults”), corresponding to the age classes of reported case notification data. Each age-class included 6 compartments representing different states of infection (Figure 1, Eq. 1 – 6). All host transition rates were simulated via a stochastic binomial probability, either sourced from the primary literature or estimated (Supp. Table 1).

**Table 1.** Model performance indicators from ABC-SMC fitting on all eight mobility models. A well-fit model has run for more generations, has a lower acceptance rate, and a higher effective sample size.

| Mobility Model | Generation | Acceptance Rate | Effective Sample Size |
| --- | --- | --- | --- |
| No Mobility | 9 | 0.0176 | 517.0201 |
| Full Mobility | 4 | 0.0131 | 856.3554 |
| Exp. Decay - Euclidean | 9 | 0.0141 | 284.9417 |
| Gravity - Euclidean | 9 | 0.0133 | 2.1528 |
| Radiation - Euclidean | 8 | 0.0238 | 87.1749 |
| Exp. Decay - Transport | 9 | 0.0141 | 75.9356 |
| Gravity - Transport | 9 | 0.0145 | 197.2948 |
| Radiation - Transport | 8 | 0.0168 | 169.8908 |

Age-specific transition rates are denoted with a ‘*’.

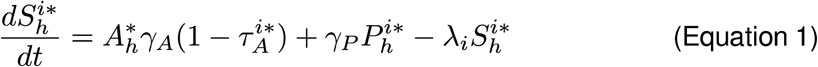

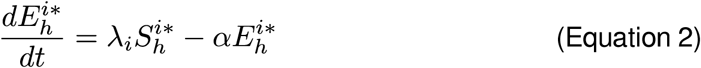

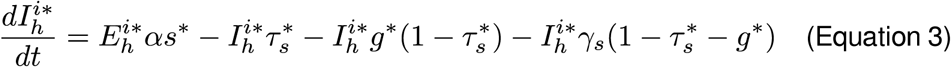

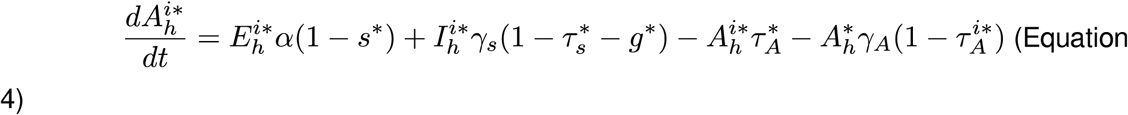

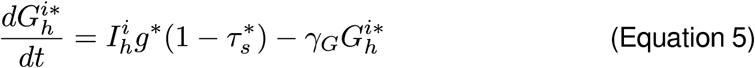

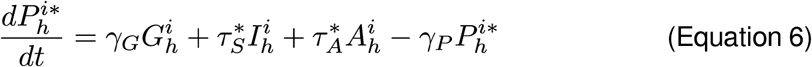

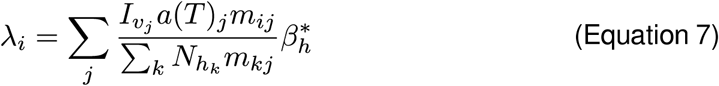

Host individuals began susceptible, *S_h_*, and transitioned to the exposed, *E_h_*, compartment via a force of infection (FOI) *λ_H_* (Eq. 1). This host FOI included the number of infectious mosquito bites received while resident and commuting and an age-specific probability of transmission, *β_h_\** (Eq. 7). Exposed individuals became either symptomatic, *I_h_*, or asymptomatic, *A_h_*, according to the symptomatic rate, *s\**, and the latent period of malaria, *1/α* (Eq. 2-4). Symptomatic individuals developed severe malaria, *G_h_,* with probability *g*,* were treated with probability *τ_s_* and entered t, or remained untreated and eventually developed an asymptomatic infection at a rate of *ɣ_s_* (Eq. 3). The treatment rates of symptomatic individuals were age-specific and based on reported treatment rates from the district of Ifanadiana (Evans et al. 2023). Asymptomatic individuals were either treated with probability *τ_A_\** or experienced waning immunity and became susceptible again following the patent period, 1/*ɣ_A_* (Eg. 4). Asymptomatic individuals were treated at much lower rates than symptomatic individuals, assuming asymptomatic cases were identified during consulations for other febrile illnesses (Supp. Table 1). Individuals with severe malaria entered the treated compartment after a severe clinical period, 1/ *ɣ_G_* (Eq. 5). Treated individuals became susceptible again at a rate of *ɣ_P_* (Eq. 6). Population sizes and proportions were held constant throughout the simulations.

The mosquito vector was modeled via 4 compartments: larval stages (L_V_), susceptible adults (S_V_), exposed adults (E_V_), and infected adults (I_V_) for each patch *i* (Figure 1, Eq. 8-11). Larvae were born as a function of the total adult population *N_V_* and temperature-dependent fecundity rates, *b(T)* (Eq. 8). Larvae experienced temperature, rainfall, and density-dependent mortality, *μ_L_(T,R,W)* (Eq. 13), as described below, and developed into susceptible adults (S_V_) via a temperature-dependent larval development rate, *ε(T)* (Eq. 8). Susceptible adults became exposed (E_V_) via the vector FOI, *λ_V_*. This was a function of the number of infected human individuals (resident and commuting) that vectors came into contact with, accounting for hosts’ individual infectiousness as a function of age and infection status, β *, and the temperature dependent bite rate, *a(T)* (Eq. 12). Exposed adults became infected (I_V_) following a temperature-dependent parasite development rate,*Δ(T),* (Eq. 10), and infected adults remained infected until death (Eq. 11). All adult mosquitoes experienced temperature-dependent mortality (*μ_A_(T))*. Vector populations were modeled at the patch-level, with no movement of individuals between patches. Transition rates were deterministic, due to computational resource limitations.

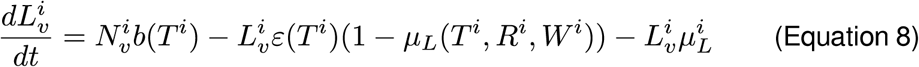

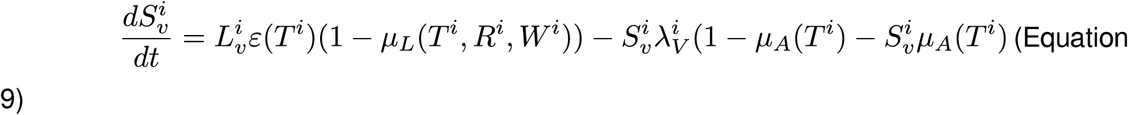

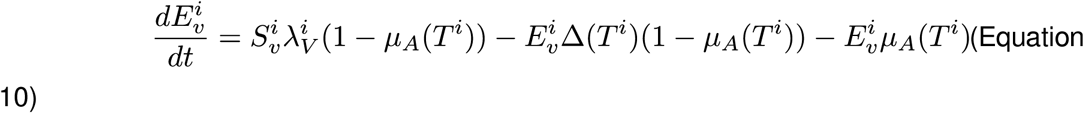

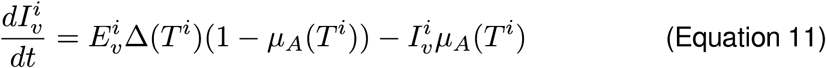

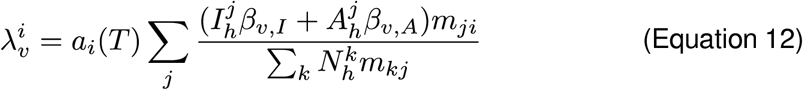

### Environmental Forcing Models

Environmental forcing of vector traits was included via patch-specific, daily measures of temperature, rainfall, and rice field flooding dynamics (Figure. 1B). Both the *Anopheles* vector and *P. falciparum* pathogen exhibit temperature-dependent life history traits (Shapiro et al. 2017). We therefore estimated daily, patch-level, temperature-dependent parameters for the vector bite rate, the parasite development rate, the vector development rate, the vector fecundity rate, the adult vector survival rate, and the larval vector survival rate. Temperature-dependent traits were estimated following either a Brier or quadratic function of temperature following Mordecai et al. (2013).

Larval mortality (𝜇*_L_)* was the sum of the temperature-dependent larval mortality rate, larval mortality due to rainfall-caused flushing events, and density-dependent mortality as a function of available larval habitat in flooded rice fields (Eq. 13-16), estimated on a daily time step for each patch. Temperature-dependent larval mortality followed Mordecai et al. (2013) (Eq. 14), where T is the daily mean temperature in that patch in Celsius. Larval mortality due to rainfall-caused flushing was an exponential function of the daily rainfall rate (Eq.15) (Tompkins and Ermert 2013). *L_f_* corresponded to the fractional larval growth state (ranging from 0-1), *K_flush_* was the maximum mortality rate due to flushing, R was the daily rainfall in mm, and 𝜏_flush_ was a scaling parameter to define the shape of the relationship between rainfall and larval mortality.

Given that rice fields are a major larval habitat for *Anopheles* in the district (Randriamihaja et al. 2025), we included density-dependent mortality rates, 𝜇*_L_(W)*, for larvae as a function of the amount of flooded rice fields in each fokontany (Eq. 16). *W* represented the amount of flooded rice field in m^2^ per fokontany and *K* was the carrying capacity of flooded rice field in individuals/m^2^.

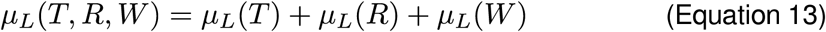

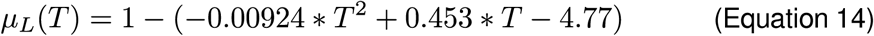

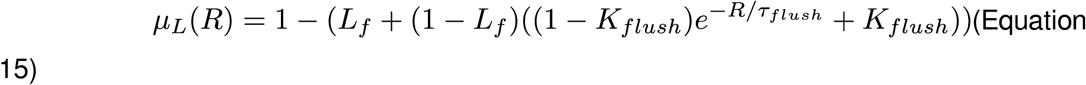

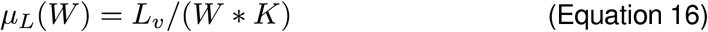

### Mobility Models

Human mobility was included via modeled daily commuting movements estimated from field-based distance networks. During the commute, hosts and vectors could infect each other in the patches hosts commuted to, defined via the mobility matrix *m* (Fig. 1C,D). However, they remained “residents” of their origin patch and did not contribute to host compartments in other patches. The mobility matrix was used to estimate the FOI on human hosts, *λ_H_*, and mosquito vectors, *λ_V,_* taking into account host commuting patterns (Eq. 7,12). These values represented the sum of the product of infectious contacts and transmission possiblilities across all commuting patches *j* for each origin patch *i*.

Mobility matrices *d* were crated from two mobility datasets that described the pairwise distance between each patch: a naive dataset representing the euclidean distance and a transport-network based distance dataset. We applied three mobility models to each dataset of pairwise distances: an exponential decay model, a gravity model that took into account the patch populations, and a radiation model that included patterns in population values of neighboring patches. For the exponential decay model, the probability of movement between patches *i* and *j* decreased with increasing distance between the patches, *d_ij_* (Eq. 17):

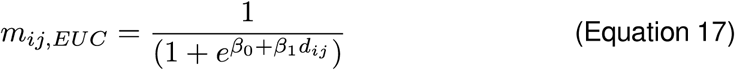

For the gravity model, the probability of movement also decreased with increasing distance, but this effect was mediated by population size, with patches with larger populations attracting more individuals from other patches (Eq. 18):

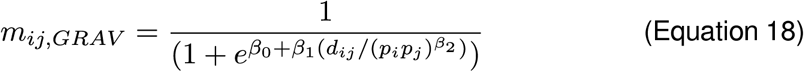

For the radiation model, the probability of commuting to patch *j* is dependent on the distance between patches *d_ij_*, the population of patches *p_i_ and p_j_*, and the total population within distance *d_ij_* of the initial patch *i (s_ij_*) (Simini et al. 2012, Kramer et al. 2016) (Eq. 19):

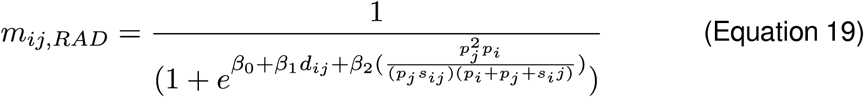

These models were applied to each mobility dataset separately. Combined with a model of no movement between patches and a “well-mixed” model where there was no limit on inter-patch movement, this resulted in eight total mobility models that were assessed. All model parameters 𝛽 were estimated via model fitting.

## Data Collection

### Malaria Case Notification and Prevalence Data

The number of monthly malaria cases per community were collected from January 2016 to June 2021 from consultation registries of all primary health centers in the District (Fig. 2). Handwritten registries were digitized and each case was geolocated to the precision of a fokontany (e.g. patch). The number of cases for children under 5 years old, children aged 5-14, and adults aged 15 years and above were summed, including both simple and severe cases.

**Figure 2.**
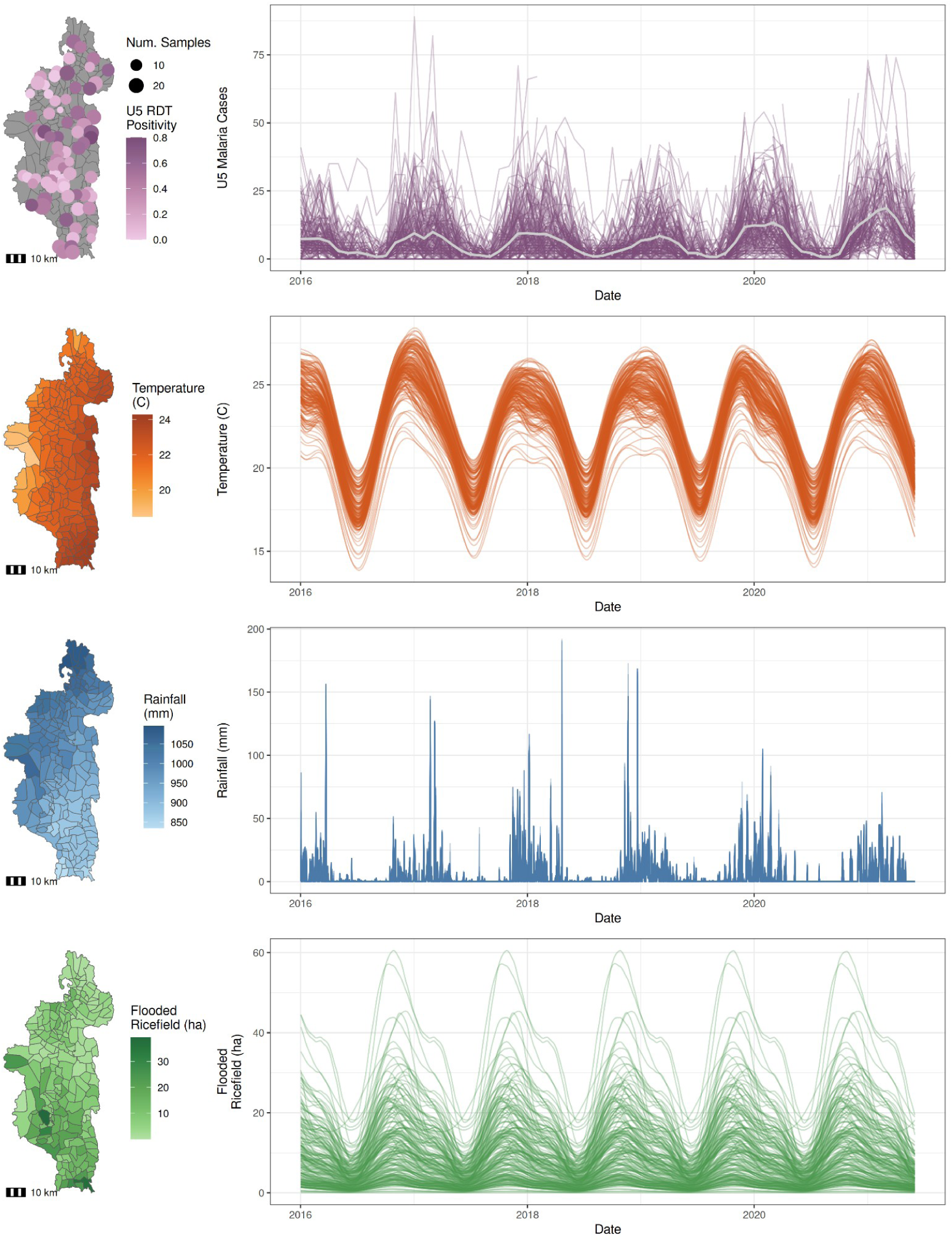
Seasonal and spatial patterns exist in malaria positivity and case rates and climatic variables. Row 1: Map of malaria positivity rates and time series of cases in children under five years old. Each individual fokontany is plotted in the time series, with the district-level mean in gray. Row 2: Average annual temperature by fokontany from 2016-2021 and daily temperature in the time series. Row 3: Average cumulative rainfall (mm) by fokontany from 2016-2021 and daily total rainfall amounts. Row 4: Average annual area of flooded rice fields (ha) by fokontany from 2016-2021 and daily flooding of rice fields.

Following Evans et al. (2025), these raw data were adjusted for underascertainment often found in passive surveillance data due to spatial biases in healthcare access using the ZERO-G method (Evans et al. 2023). The method was applied separately to each of the three age classes above to estimate a monthly case rate. Case rates were transformed into case counts by scaling by the population of each age class and were ultimately aggregated into the two age classes used in the model: children under 5 years old and persons aged 5 or older, referred to as adults.

Malaria prevalence for children under 5 years old was collected via the IHOPE survey conducted in April-June 2021 in 1600 households in 109 fokontany throughout the District (Fig. 2). Briefly, children under 15 years old who consented to the study were tested for active malaria infection using SD One Step Malaria HRP-II(P.f) and pLDH(Pan) Antigen Rapid Tests. Those who tested positive were provided with a standard treatment of artesunate amodiaquine and paracetamol, with duration and dosage in accordance with national guidelines. For this study, we limited our prevalence estimates to children under 5 years old, so that it paired with the child age class in the dynamic model, resulting in 969 samples. Patch-level prevalence was defined as the mean prevalence of all individuals in clusters within that patch.

### Distance Network Data

Mobility models were run on two distance networks: a naive network and a transport network. The naive network represented the pairwise euclidean distance between the centroid of each fokontany. The distance based on a transport network was estimated using a comprehensive dataset of over 25,000 km of tracks and footpaths and 5,000 residential zones available in OpenStreetMap (Ihantamalala et al. 2020). The shortest distance between all residential zones on the network was estimated using the Open-Source Routing Machine service (Luxen and Vetter 2011). Pairwise distances between each fokontany were then the average building-weighted distance between the residential zones of each fokontany. Both approaches resulted in a matrix of pairwise distances between all patches *i* and *j* (*d_ij_*).

### Environmental Data

As a mosquito-borne disease, malaria dynamics are highly dependent on the surrounding environment, particularly climate and the availability of larval habitat (Paaijmans et al. 2009, Hardy et al. 2013, Chan et al. 2022). Data on temperature, rainfall, and ricefield flooding dynamics were used as environmental forcing to model vector dynamics, as detailed above (Fig. 2). Temperature data was sourced from the MODIS Aqua Daily Land Surface Temperature dataset following (Evans et al. 2025). This method uses a gap-filling algorithm to create daily estimates of land temperature at a 1 km resolution (Shiff et al. 2021), which was then extracted to the fokontany-level. This resulted in a daily time series of temperature for each fokontany from January 1 2006 through December 31 2021. Daily rainfall data was derived from the Africa Rainfall Climatology 2 Model (Novella and Thiaw 2013) and the spatial mean extracted at the fokontany-level, resulting in a daily time series of rainfall for each fokontany from January 1 2006 through December 31 2021. Rice field flooding dynamics were described by estimating the daily value of the amount of flooded rice fields in m^2^ per fokontany using a field-tested, radar-based classification algorithm developed for this region of Madagascar (Randriamihaja et al. 2025). This data was collected from the Sentinel-1 satellite constellation for January 1 2016 through Dec 21 2021. The total flooded rice field area was then estimated daily at the fokontany-level. Because Sentinel-1 timeseries data was not available for dates before 2016, the average for that day of the year was used for January 1 2006 - December 31 2015.

### Parameter Estimation

Transition rates and mobility model parameters were estimated for each mobility model through Approximate Bayesian Computing based on Sequential Monte Carlo (ABC - SMC, Sisson et al. 2007). ABC-SMC allows the estimation of parameters where the log-likelihood is intractable or expensive to compute, or when multiple datasets or customized error functions are to be considered (Beaumont 2010, Minter and Retkute 2019). In our case, it enabled us to fit the model to malaria prevalence and incidence data simultaneously and include true environmental forcing data which prohibits exact estimation of the log-likelihood. Applying ABC-SMC, we fit the dynamic models to monthly case notification data for each age group from Jan 2016 - June 2021 and malaria prevalence data in children under-5 from May 2021, for a total of five summary statistics: the root mean square error between simulated and observed monthly adult cases by fokontany; the root mean square error between simulated and observed monthly child cases by fokontany; the proportion of months within 20% of the observed case numbers occurring during the malaria peak (November - May); the mean absolute error in simulated and observed prevalence in children; and the correlation of the average annual simulated and observed case count for each fokontany. The district-level mean of each statistic was calculated via a weighted mean, where the weight of each fokontany was proportional to the number of months of complete, non-zero observations in the training data.

The ABC-SMC process was run for ten generations or until the particle acceptance rate dropped below 1% for each mobility model. The initial generation began with manually selected thresholds for the summary statistics, with sequential generations adopting the median of the prior generation’s accepted particles’ performance as the threshold. Potential particles were generated from a covariance matrix of the previous generation of accepted particles containing the 25 nearest neighbors, following Minter and Retkute (2019). Each generation was simulated until 1000 particles were accepted. Posteriors were then generated from the last generation’s distribution of 1000 particles, weighted by a multivariate prior drawn based on the covariance matrix of the 25 nearest neighbors for that particle.

### Simulating Spatially Targeted Interventions

#### Intervention Scenarios

The best fit mobility model was then used to simulate the impact of spatially targeted and non-targeted interventions in the district. Three interventions were considered: active case detection (ACD), long-lasting insecticide net (LLIN) distributions, and indoor residual spraying (IRS) campaigns. All three interventions are part of the Strategic Plan of the Madagascar National Malaria Control Programme (*Programme Nationale de Luttre Contre le Paludisme, PNLP)* for the period of 2023-2027. Interventions were simulated beginning on January 1 2016.

ACD interventions were simulated to increase treatment rates for symptomatic individuals across all age groups, τ*. Base treatment rates were estimated for children under-5 years old and adults from the 2021 IHOPE survey, with rates of 0.5658 and 0.3319, respectively. The equally distributed intervention increased these rates by 50% to 0.8486 and 0.4979 for children and adults, respectively, across all fokontany in the district. The targeted interventions increased both treatment rates to 1 in the targeted fokontany. This corresponds to the PNLP’s malaria treatment rate goal for 2026. To account for gradual increases in treatment rates, all interventions increased the treatment rate linearly from the baseline treatment rate to the goal treatment rate over the six month period prior to the start of the intervention.

LLIN distribution interventions impacted adult mosquito bite rates and mortality rates.

Mosquito bite rates were reduced as a function of use rates of LLIN *U_LLIN_*, distribution coverage rates *C_LLIN_*, and mosquito endophagy rates, *x*. The bite rate when LLINs were present was:

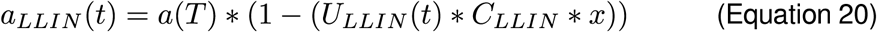

LLIN use rates were described via a time-dependent decay function of LLIN usability and condition, assumed to be a 0.25 reduction in proper use per year (Randriamaherijaona et al. 2017b):

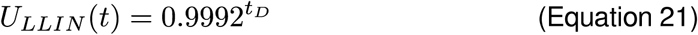

where t_D_ is the time since mosquito net distribution. The mosquito endophagy rate, *x*, was assumed to be 0.3 (Ratovonjato et al. 2014), ensuring LLIN interventions only impacted indoor biting mosquitoes.

Mosquito mortality rates were increased by LLIN interventions as a function of the bite rate with LLIN and the permethrin-induced mortality rate *μ_p_*. The total adult mortality rate when LLINs were present was:

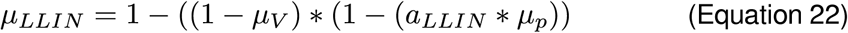

Permethrin-induced mortality decayed over time from distribution *t_D_* following a bioefficacy decay rate of permethrin estimated from field-data from Madagascar (Randriamaherijaona et al. 2017a)

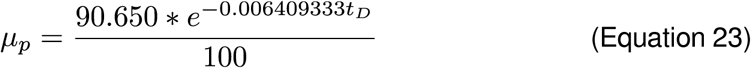

LLIN distributions were set to occur on August 1st on a three year cycle beginning in 2003, following PNLP policy. The baseline coverage rate was set at 50%, based on IHOPE 2021 data, and simulated interventions increased these coverage rates. Equally distributed coverage rates increased existing coverage rates by 50% to equal 75% coverage in all fokontany and spatially-targeted interventions increased coverage rates by 100% to equal100% coverage in targeted fokontany, to keep the district-wide effort similar between targeted and non-targeted interventions.

### IRS campaigns impacted adult mosquito mortality rates, accounting for the waning bioefficacy of bendiocarbs following Sherrard-Smith et al. (2022). Mortality with IRS present, μ_IRS_, therefore was

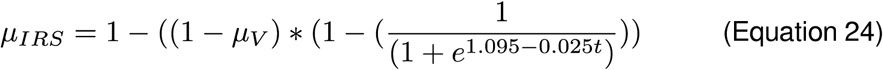

IRS campaigns were set to occur on November 1 of each year on an annual basis beginning in 2016. Baseline coverage rates were 0, as no IRS campaigns are currently done in Ifanadiana. Equally distributed coverage rates increased IRS coverage to 0.30 in all fokontany, following PNLP targets. Spatially-targeted interventions increased IRS coverage rates to 0.60 in targeted fokontany. This difference in coverage rates was done to impose similar district-wide efforts for both targeted and non-targeted interventions.

### Spatial Targeting Scenarios

Five scenarios of spatial targeting were considered in this study. All targeted scenarios targeted 50% of the fokontany with approximately twice the coverage rate of a non-targeted intervention, so that the overall district-wide intervention effort remained similar between non-targeted and targeted interventions. The first scenario targeted those fokontany nearest to health clinics on the road network, representing the least isolated and generally more populated fokontany. The second scenario targeted those fokontany further from health clinics on the road network, representing more isolated fokontany with lower access to health care.

The third scenario targeted the fokontany with the highest malaria incidence rates in 2016, representing fokontany with potentially higher malaria exposure. The fourth scenario targeted the most central fokontany on the radiation mobility network, representing the most “connected” fokontany. This was defined as the average probability of movement to each patch from other patches. The fifth scenario was non-targeted, and simply applied the intervention to all fokontany equally at the lower coverage rate.

### Assessing and Visualizing Intervention Scenarios for Decision-Making

Each combination of intervention and spatial targeting scenarios (15 total) was simulated 49 times using the best fit parameters from the model fitting exercise. We then estimated the median number of simple and severe malaria cases averted per age-class relative to the baseline scenario with no additional interventions to assess the impact of each intervention scenario. We also assessed combining two or all three interventions for each of the five targeting scenarios at low and high levels of coverage. Low coverage was equal to 1.5x base levels, with a maximum of 100% coverage. High coverage was equal to 2x base levels, with a maximum of 100% coverage. Coverage was increased in targeted fokontany, with other fokontany retaining base coverage levels.

While coverage rates were set lower for non-targeted scenarios to approximate equal intervention efforts at a district-wide level for targeted and non-targeted interventions based on the number of patches targeted, the total resources required by an intervention can also depend on the distance needed to travel to implement the intervention and the number of households impacted. We therefore estimated the total distance from targeted fokontany to the nearest health center, the total distance from targeted fokontany to the district hospital, the total number of households targeted by IRS and LLIN, and the total number of individuals benefiting from ACD for each targeting scenario to better estimate the overall resources required for each scenario.

As part of this model health district’s program to conduct actionable research, we developed an accompanying web application to allow local health actors to explore a wider range of intervention scenarios. This includes the ability to choose single or multiple interventions, the type of spatial targeting, and the coverage rate of the intervention. Users can then explore time series and maps of the number of simple or severe malaria cases resulting from the simulated intervention scenario at fokontany and district-levels. This application was developed using R Shiny and is available at https://smaller.pivot-dashboard.org/ on the ‘Interventions’ tab.

## RESULTS

### Spatial Variation in Parameters due to Environmental Forcing

There was a range of spatial heterogeneity in the environmental forcing parameters. When considering daily variation (e.g. spatial differences at the same time point) across the district, temperature varied by 5.15 - 6.95 C, rainfall by 0 - 281.3 mm, and flooded ricefields by 18.6 - 60.5 ha, depending on the season (Figure 2). There was large daily variation in temperature-dependent vector parameters throughout the study period (Supp. Table 2, Supp. Figure 2). Pathogen and larval development rates were the most varied temperature-dependent rates across space with average coefficients of variation across the district of 0.156 (0.0813 *sd)* and 0.155 (0.0817 *sd)*, respectively. This was equivalent to certain patches having development rates over twice as high as the average development rate during the same timestep. In contrast, adult survival was least sensitive to spatial heterogeneity in temperature, with rates never differing by more than 0.06 across the district (Supp. Table 2). Larval mortality due to rice-field carrying capacity also exhibited distinct spatial heterogeneity, with an average coefficient of variation above 0.83 (Supp. Table 2). The median range in maximum larval carrying capacity across the district was 9.6 million larvae per fokontany, with certain fokontany experiencing carrying capacities over five times higher than the average carrying capacity during the same time step. Rainfall induced larval mortality rates, which were based on coarser resolution rainfall data and included a non-linear functional response, were notably less varied across space (Supp. Figure 2, Supp. Table 2). The median difference in rainfall-induced mortality rates across the district was 0.00, with only 5.0% of timesteps with differences greater than 0.1.

**Table 2.**
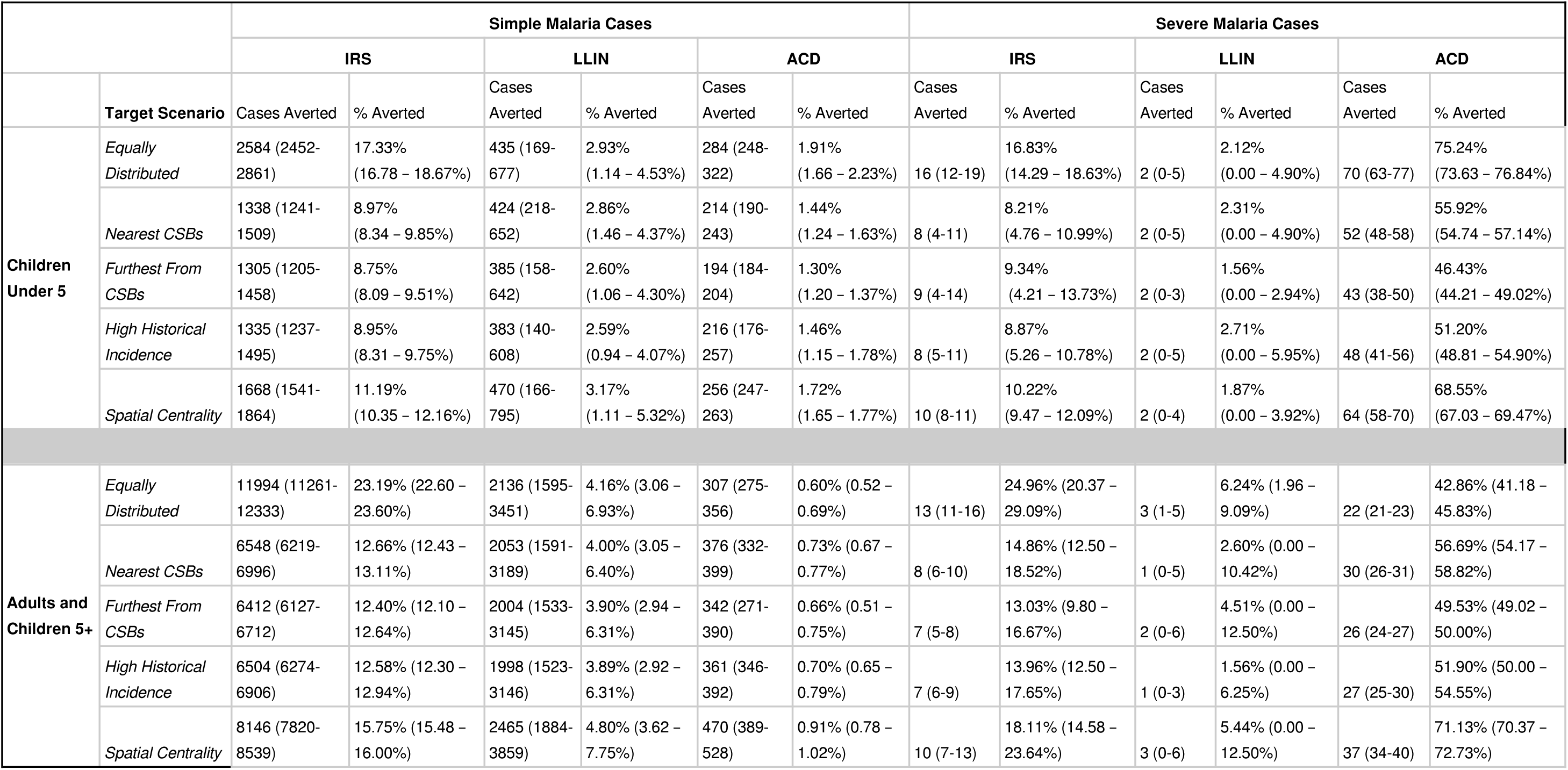
Equally distributed scenarios generally avert the most cases for all three types of interventions, particularly for children under 5. Average annual cases averted by targeting scenario and intervention for simple and severe malaria cases for children under 5 years old and adults and children 5 years and older. The range of annual averted cases between 2016-2021 are shown in parentheses. ACD: active case detection; IRS: indoor residual spraying; LLIN: long-lasting insecticidal nets.

## Model Performance

The majority of mobility models reached convergence with an acceptance rate higher than 1% within 8 or 9 generations of the adaptive threshold, with the exception of the Full Mobility model which reached extremely low acceptance rate values after 4 generations (Table 1). Both the No and Full Mobility models had relatively high effective sample sizes (ESS) over 500, representing the effective number of independent samples in the final particle set. Higher ESS is associated with higher precision and convergence among the MCMC sampling algorithm for a model. Among the more complex mobility models, on the euclidean distance network, only the exponential decay model had an ESS value over 100, while both the gravity and radiation models neared 200 ESS for the transport network (Table 1). We evaluated performance on 499 simulations drawn from the posteriors for each mobility model. The No and Full Mobility models performed the poorest across all five summary statistics, with the exception of RMSE - U5 Cases, where the No Mobility model performed similarly to the network-based models (Figure 3). Among the six distance-based models, there were no consistent trends in performance across the five summary statistics and all six performed similarly (Figure 3). To identify the best fit model, we ranked the models within each summary statistic according to their median summary score and estimated the mean ranking by model. The exponential decay model using network distance from transport data (Exp. Decay - Transport model) was identified as the best fit and used to compare interventions.

**Figure 3.**
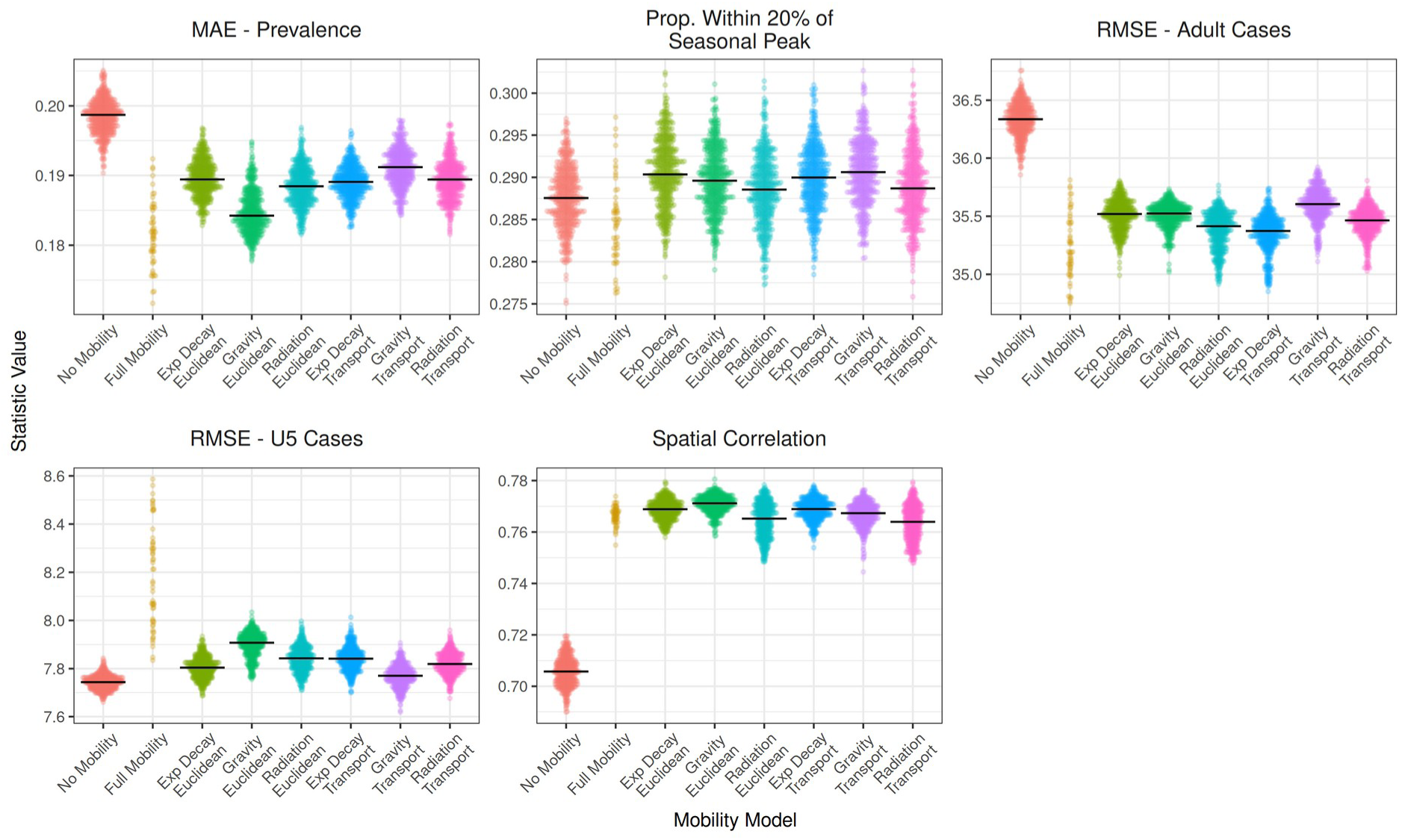
Mobility models that included patch-specific movement performed similarly on summary statistics. Results of 499 individual simulations from the posterior for each mobility model (points) with the median value represented by the bold line. Simulations with performance scores outside of the initial threshold values and the median value of the Full Mobility model were removed to aid in visualization.

### Intervention Scenarios

When Equally Distributed without a targeting scenario, IRS interventions were the most effective against simple malaria cases, averting on average 23.2% of cases in adults and 17.3% of cases in children (Table 2). LLIN distributions and active case detection were considerably less effective, reducing cases between 2.93 - 4.16% and 0.06 - 1.96%, respectively (Table 2). However, when considering severe malaria cases, ACD was the most effective intervention, averting on average 42.9% of adult cases and 75.2% of childhood cases (Table 2). The other interventions performed similarly for simple and severe cases, although there was more uncertainty around severe cases averted given the lower overall numbers (Table 2). LLIN distributions, in particular, had very little impact on severe malaria cases (Figure 4, Table 2)

**Figure 4.**
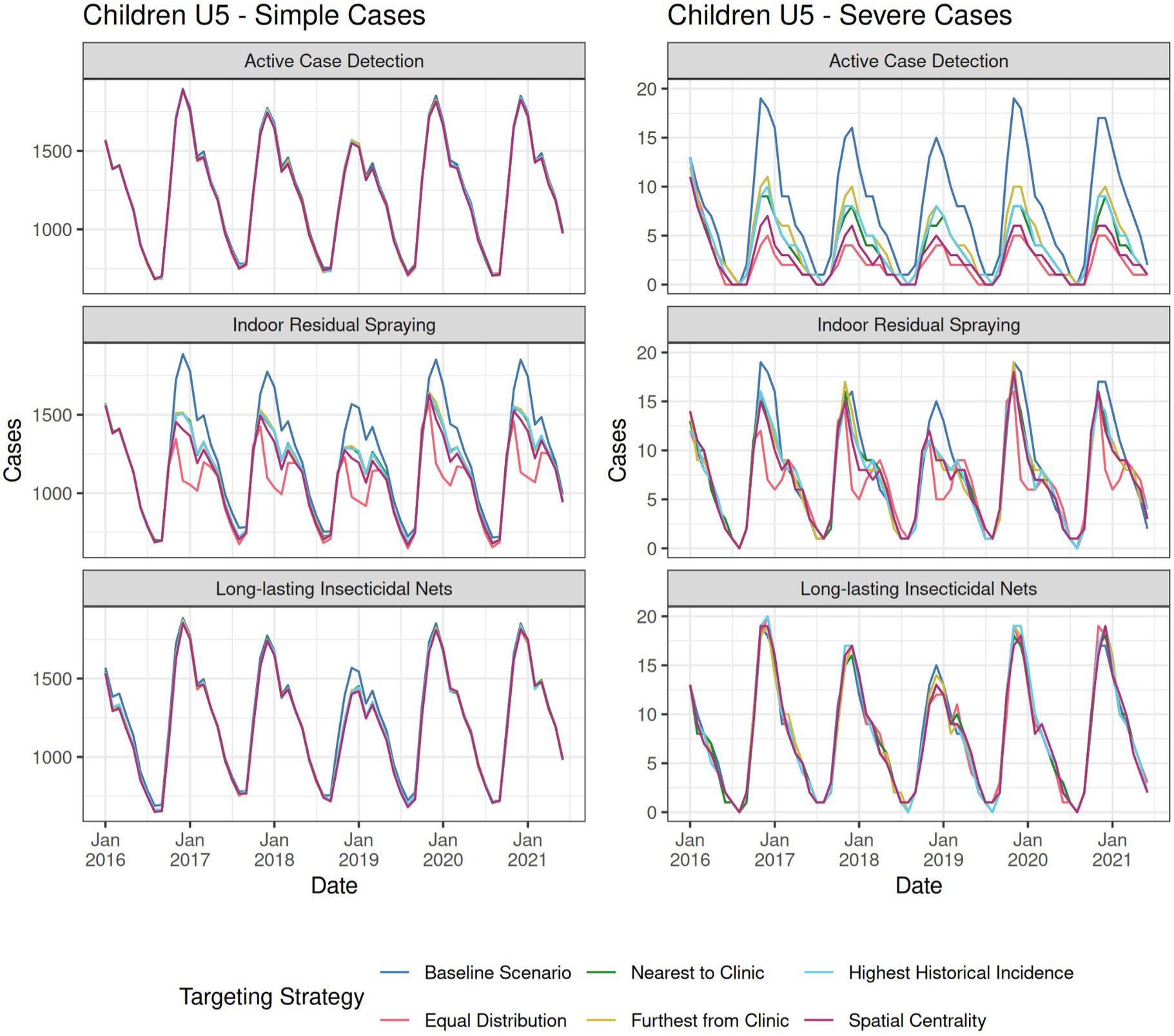
Equal Distribution and Spatial Centrality targeting strategies tended to perform the best for all three interventions. Each line represents the median of 499 simulations of the number of cases for each combination of targeting strategy (line color) and intervention (row) for simple (left) and severe (right) malaria cases in children under 5 years old. Simulations are based on draws from the posterior of the best fit model. Adults are shown in Supplemental Figure 3.

In general, targeting scenarios based on the distance to the health clinic or historical incidence performed worse than the Equally Distributed scenario for all interventions and case types (Figure 4, Table 2). However, the Spatial Centrality targeting scenario performed better than or equal to the Equally Distributed scenario for the LLIN and ACD interventions, and was the second best targeting scenario for IRS (Figure 4, Table 2). This effect was diminished for severe cases, where the Equally Distributed scenario consistently performed best, except for adults with ACD, where the Equally Distributed Scenario averted the fewest number of cases (Table 2).

When considering the resources required for each intervention, a similar number of houses were targeted for all IRS scenarios and similar sized populations were reached by all ACD campaigns (Table 3). However, the Equally Distributed scenario had the highest resource use with regards to distance to the nearest health clinic or district hospital. It included distances to clinics over three times that of the Nearest to Health Clinic scenario, which had the smallest total distance (Table 3). The Spatial Centrality scenario resulted in the largest population covered by ACD, covering over 25,000 more individuals than the Equally Distributed scenario, while maintaining the lowest overall distance to the district hospital (Table 3). There was little difference in the number of houses included in IRS across all five scenarios (Table 3), while the number of houses included in LLIN campaigns ranged from 17504 to 28807 depending on the scenario (Table 3).

**Table 3.** The equally distributed targeting scenario had the highest transport needs, while covering similar numbers of households and populations as other interventions. An estimation of resources required for each spatial targeting scenario based on the travel distances needed to implement the intervention, the number of households covered by the intervention, and the total population covered by the intervention.

|  | Total Distance from<br>Clinic (km) | Total Distance from<br>District Hospital (km) | Houses Targeted for<br>IRS | Houses Targeted for<br>LLIN | Population Covered<br>by ACD |
| --- | --- | --- | --- | --- | --- |
| Equally Distributed | 1375.22 | 12514.99 | 11523 | 28807 | 126848 |
| Nearest to Clinic | 368.56 | 6545.26 | 12508 | 20847 | 125881 |
| Furthest from Clinic | 1006.66 | 5969.72 | 10537 | 17562 | 106004 |
| Historical Incidence | 615.15 | 6468.6 | 11333 | 18889 | 115562 |
| Spatial Centrality | 585.61 | 4812.27 | 13698 | 22830 | 155260 |

We evaluated different coverage levels and combinations of interventions to identify scenarios that could be used by stakeholders to eliminate malaria in the District. The most effective scenario at averting simple cases was a combination of all three interventions at high coverage using an Equally Distributed targeting strategy, which averted 43.69% of adult cases and 37.04% of childhood cases, on average (Supp Table 2). The Equally Distributed targeting scenario continued to perform best, outperforming other targeting scenarios at all levels of coverage and intervention combinations (Supp. Figure 4, Supp Table 2). With regards to severe cases, any scenario that included ACD at high coverage levels Equally Distributed across the District was able to avert 100% of severe cases for both age classes (Supp. Figure 4,5). Those scenarios using Spatial Centrality targeting performed second best, averting between 67.52 - 75.16% of cases when including ACD at high coverage levels, while there was little difference between the remaining three targeting scenarios (Supp. Table 2).

**Figure 5.**
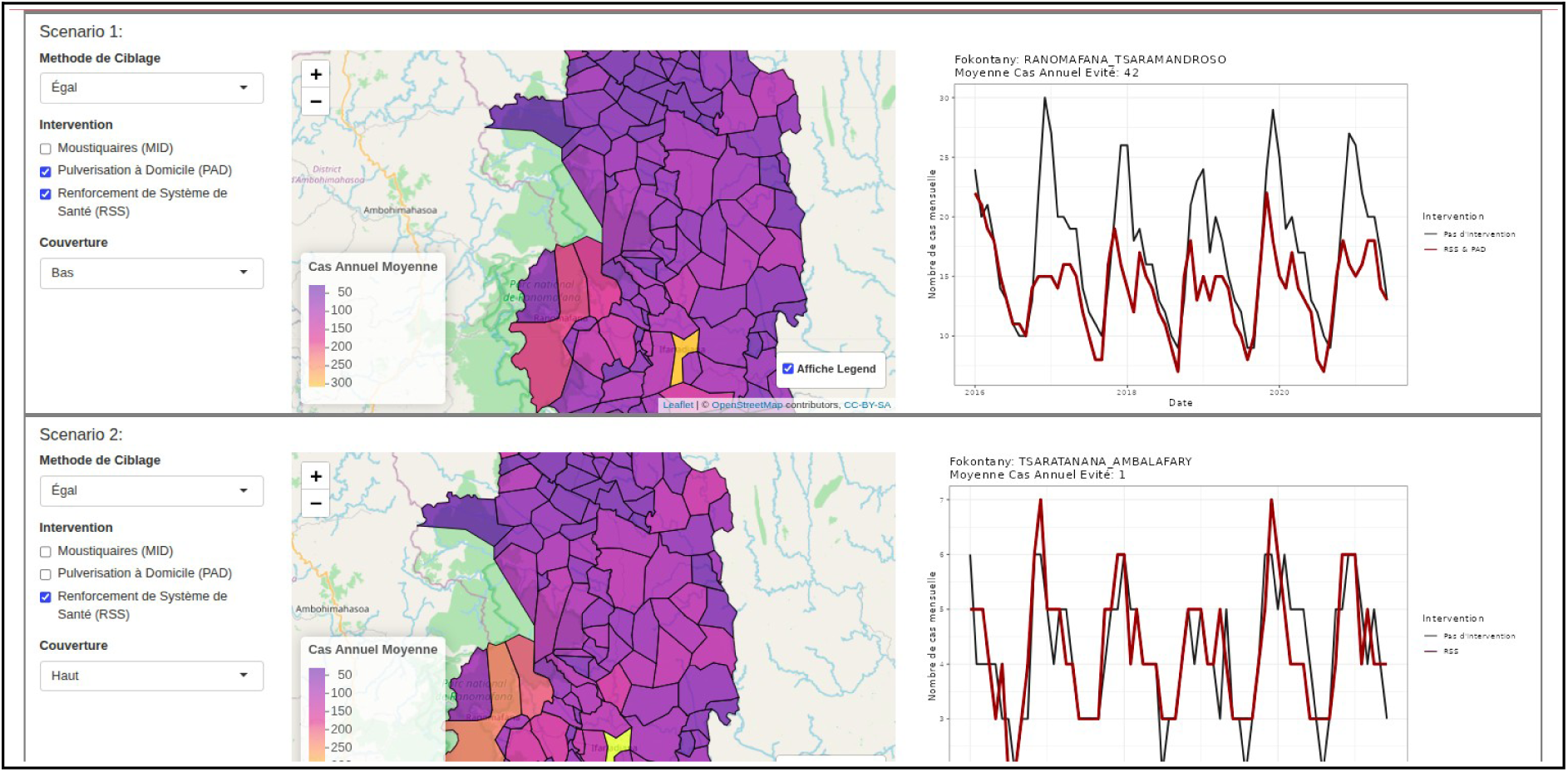
Screenshot of web application displaying the results of different scenarios of combinations of interventions and spatial targeting. The application is accessible at https://smaller.pivot-dashboard.org/ on the Interventions tab.

An accompanying web application displays the results of intervention scenarios at fokontany and district levels (Fig. 5). Users can choose which combination of interventions, spatial targeting scenario, and coverage level to consider and evaluate the resulting impact on simple and severe cases compared to a base scenario. It also displays spatial patterns in the number of cases averted. These functionalities were part of a larger effort to inform local decision making in malaria control by combining an early warning system (Evans et al. 2025) with scenarios of intervention impact into the same application.

## DISCUSSION

As malaria control progress slows and funding decreases, spatially targeted interventions offer a potential solution to distribute limited resources more efficiently. However, evidence regarding the effectiveness of spatially-targeted interventions is mixed, and dependent on the spatial scale of the targeting. Designing spatially-targeted control programs within a health district could allow for local tailoring of national-level interventions and result in more effective interventions. We developed a spatially-structured dynamic model of malaria for a rural district in southeastern Madagascar to compare different spatial targeting strategies of three common malaria interventions conducted at the community-scale. Notably, the model was based on empirical environmental data and distance networks and fit to field-derived prevalence and incidence data at the community-scale. Our modeling exercise illustrates the importance of including spatial heterogeneity in the modeling of malaria at the community-scale, particularly with regards to environmental forcing. We find that Equally Distributed interventions tend to perform better than or equally as well as spatially targeted interventions across all combinations of interventions in this high transmission setting. However, spatial targeting that takes into account the spatial distribution of community catchments and their populations can be as effective as equally distributed interventions in certain cases while mobilizing fewer resources. This suggests that effective tailoring of interventions should use spatial structure to guide campaigns, rather than hotspot identification.

Our results demonstrate the importance of including spatial structure in the environmental forcing variables of a dynamic malaria model. This was particularly true for vector and pathogen traits that were temperature dependent, given the wide-range of temperatures seen across the District and the large number of traits that were temperature sensitive. This agrees with prior work identifying temperature as a strong driver of spatial heterogeneity in malaria transmission through both mechanistic ((Mordecai et al. 2013, Shapiro et al. 2017) and statistical models (Ryan et al. 2015, Weiss et al. 2019, Whittaker et al. 2022). In addition, the temperature range seen across the district (15-27 C) corresponds with the range with the highest amount of variation and uncertainty in thermal response curves for *Anopheles* mosquitoes and malaria (Johnson et al. 2015). Rainfall and flooding data were available at coarser resolutions and the traits dependent on these variables were less sensitive to spatial variation across the range seen in the District (Supp. Figure 2). Although commonly incorporated into statistical models (e.g. Wimberly et al. 2022, Pourtois et al. 2023, Villena et al. 2024)), few mechanistic models include multiple environmental forcing variables at once (Eikenberry and Gumel 2018, but see Tompkins and Ermert 2013, Okuneye and Gumel 2017). This has been applied even more rarely at local, community scales, where climate data may be limited (Skinner et al. 2024), although satellite-based climate data and models, such as ERA5 or MODIS, are becoming more easily accessible. Given the increasing availability of this data, future work modeling malaria transmission via dynamical or mathematical models should include spatially-explicit environmental forcing dynamics when possible.

We found that including realistic spatial structure in human mobility increased model performance, regardless of whether this structure was based on a naive model of Euclidean distance or a more complex model that includes true road networks. This is in contrast to prior work that found that including true road networks resulted in better estimates of geographic barriers to care than euclidean distances (Shahid et al. 2009, Garchitorena et al. 2021, Ihantamalala et al. 2021). However, distance in our model was scaled to range from 0 - 1, and not a direct calculation of travel time based on the distance. As suggested by established theory (Love and Morris 1972), the mobility matrices based on both datasets were therefore highly correlated and resulted in similar mobility and malaria dynamics. This suggests that the simpler euclidean distance network can be used when a true road network is not available and absolute distances or travel times are not necessary for the mobility model. However, our networks did not include information on road condition or type, which could greatly impact travel time. Including such information would likely reduce the correlation between the two datasets and provide more accurate estimates of perceived distance.

In spite of the importance of spatial structure to the dynamical model, spatially naive interventions, which were applied equally across the district, tended to be the most effective. While much effort has been put into the identification of malaria “hotspots” to guide targeting, evidence for the effectiveness of spatially targeted interventions is mixed (Stresman et al. 2019, Khundi et al. 2021). Spatial targeting has proven useful in malaria elimination zones, where individual cases or clusters of cases may act as a source for an outbreak (e.g. the Zanzibar Malaria Elimination Programme, Björkman et al. 2019), but may be less useful in areas with heterogeneous, but overall high, rates of malaria transmission, where these hotspots may not lead to increased malaria transmission in surrounding areas (Bousema et al. 2012). Given the high case rates of malaria seen in Ifanadiana (over 100 cases per 1000 population annually, Hyde et al. 2021), it is likely that overall transmission rates are too high for hotspot targeting to be effective, even at the community-scale. However, it is notable that the best performing spatially targeted intervention was not based on case incidence or prevalence, as is often used in hotspot targeting, but on the underlying spatial structure of the transport network. This suggests that incorporating spatial patterns in population density and movement when allocating resources could be more effective than guiding interventions using historical disease case rates, if spatial targeting is to be used.

Concerning the ideal community-scale intervention scenario for this region of Madagascar, our results suggest that interventions that target the vector itself, particularly IRS, are most effective at controlling simple malaria cases. The performance of IRS relative to LLIN campaigns is likely due to some assumptions of our model regarding LLIN effectiveness in Madagascar. Following field data in Madagascar, we assumed mosquitoes were 30% endophilic (Ratovonjato et al. 2014), with the majority of biting occurring outside the home where LLINs provide little to no protection (Briët and Chitnis 2013). We also included a waning pesticide efficacy and maximum rate of correct use and pesticide-induced mortality that limited the effectiveness of LLINs. Finally, because LLIN coverage was already at 50% in the district, it could not be increased in simulations as much as IRS, which is currently not practiced in the district. Increased returns on LLIN-based interventions may be more likely by focusing on increasing use rates themselves, rather than simply increasing the coverage of distribution campaigns, which had limited returns at very high values. Indeed, the decreasing ability of LLINs to control malaria due to changing mosquito behavior and insecticide resistance has been noted globally (Okumu 2020). With regards to resource use, the Equally Distributed scenarios required the most travel distance from clinics and the district hospital, a major source of costs in IRS and LLIN campaigns. The Spatial Centrality scenario required about a third of the travel distance of the Equally Distributed scenario, and was able to cover a similar amount of households. Decision makers should therefore take into account the trade-off between the effectiveness of district-wide interventions and their potential costs, depending on their budget and requirements. None of the interventions were able to fully eliminate malaria cases, although ACD was able to avert 100% of severe malaria cases under a “perfect” detection scenario. Madagascar’s current strategy aims to increase case detection and treatment rates by expanding malaria community case management to all ages and allowing proactive case detection by CHWs and other health teams. Our simulation results support this as an effective intervention given the high rates of endemicity, agreeing with pilot field studies that tested these interventions in neighboring regions of Madagascar (Ratovoson et al. 2022, Garchitorena et al. 2024).

While our dynamic model was complex, it necessarily simplified several aspects of malaria transmission dynamics. We did not include individual-level immunity nor model the actual parasite within human hosts, both of which have been shown to be important for malaria transmission models (The malERA Refresh Consultative Panel on Combination Interventions and Modelling 2017, Smith et al. 2018). In particular, immunity can interact with environmental forcing to drive malaria seasonality in endemic regions such as Ifanadiana (Childs and Boots 2010, Laneri et al. 2015). We instead included these dynamics using a simplified two-class age model with differing rates for adults and children and allowing for an age-specific waning immunity rate, given the computational limitations of explicitly modeling 195 patches. The model also did not include spatial structure in existing interventions, particularly access to care and LLIN use, which exhibit strong spatial heterogeneity across the District (Garchitorena et al. 2018). These behaviors may change over time in unpredictable ways in response to perceived risk (Pinchoff et al. 2015). We therefore chose to use a District-level mean for the existing intervention coverage rates, based on field-collected data. This could potentially influence our findings regarding the spatially-targeted interventions if, for example, some patches already have very high or low levels of coverage. However, given the clear superiority of Equally Distributed interventions, we expect this difference would primarily be qualitative and patch-specific.

Our study agrees with prior work that finds that equally distributed interventions are the most effective at averting malaria cases in endemic regions. However, we also find that they are the most costly, particularly with regards to travel and transport. Spatially targeted interventions, particularly those based on spaital structure and not historical incidence, were nearly as effective as equally distributed interventions at half the travel costs. Incorporating the underlying spatial structure of the population could be a more efficient use of resources when they are limited, and should be explored in other contexts.

## Data Availability

All code and data needed to run model simulations is available on Figshare.

## ACKNOWLEDGEMENTS

We would like to thank the health care workers of Ifanadiana District for their dedication to treating patients and collecting high quality data. We would like to acknowledge the work of Ann Miller and the research team at INSTAT, who implement the IHOPE survey.

## AUTHOR CONTRIBUTIONS

MVE, BR, and MHB, AG conceptualized the study. MVE, BR, VH, CR, TC, FAI, MR, AG contributed to designing the methodologies concerning data collection and analysis. MVE, FAI, MR, and AG collected the data and MVE, BR, FAI, MR and AG conducted analyses and all authors interpreted results. Project supervision was provided by BR, MHB, KEF, FAI, OR and AG. The manuscript was drafted by MVE, BR, and AG and all authors contributed critically to the drafts and gave final approval for publication.

