## Supplemental Figures for "Leveraging spatial structure to design spatially-targeted malaria interventions at the community-scale"

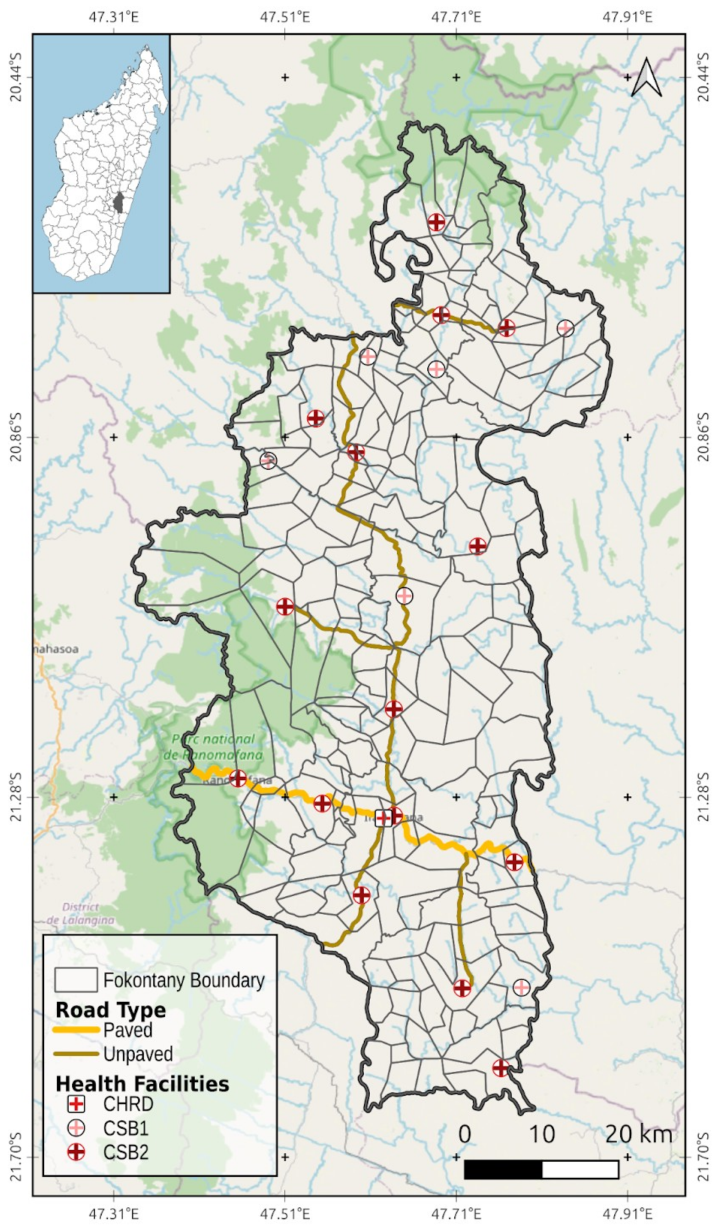


**Supplemental Figure 1.** Map of study area of Ifanadiana District, Vatovavy, Madagascar with primary transport network and health clinics. Inset map shows district's location within Madagascar. Basemap Source; OpenStreetMap

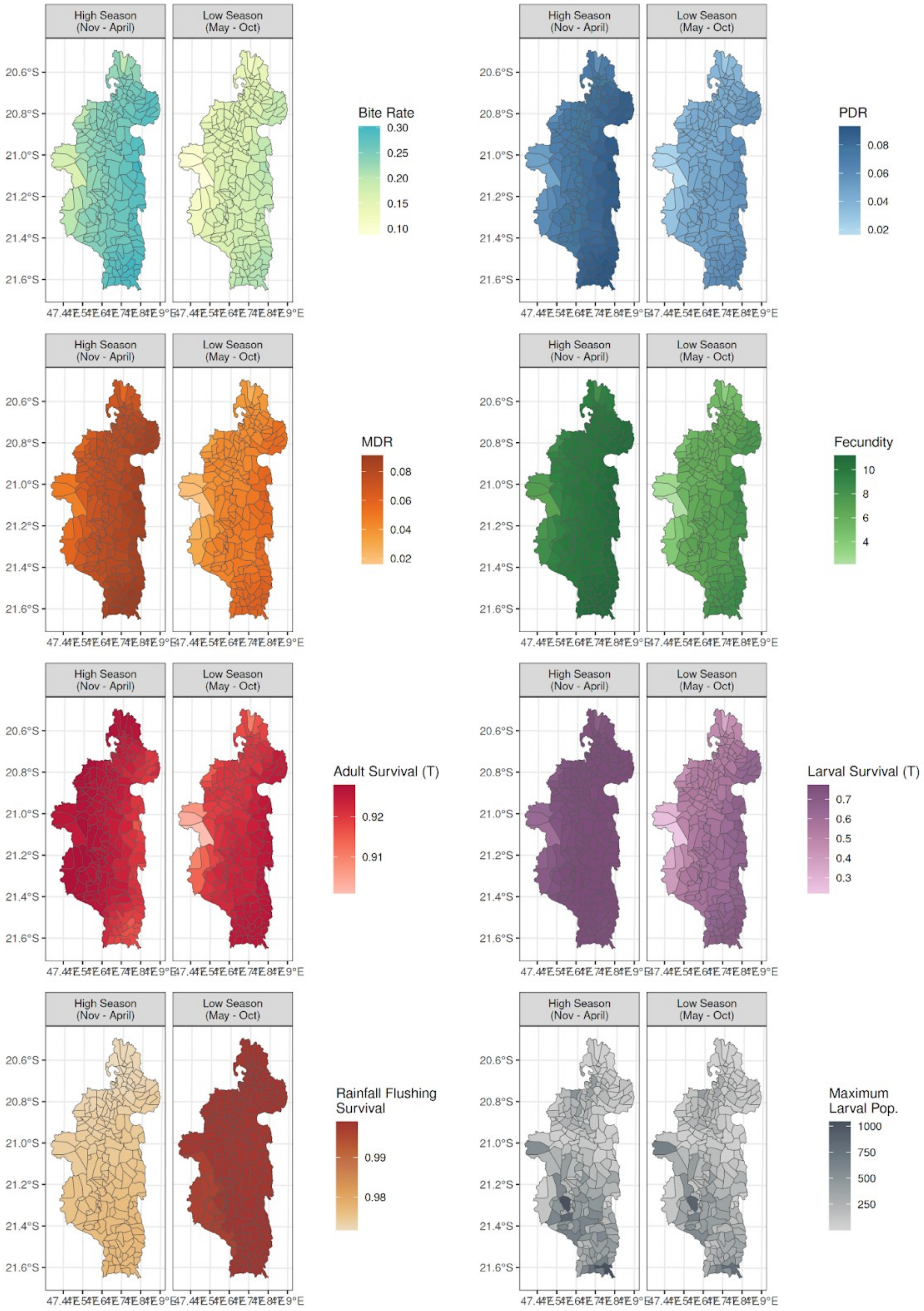


**Supplemental Figure 2.** Variation in environmental forcing by malaria season

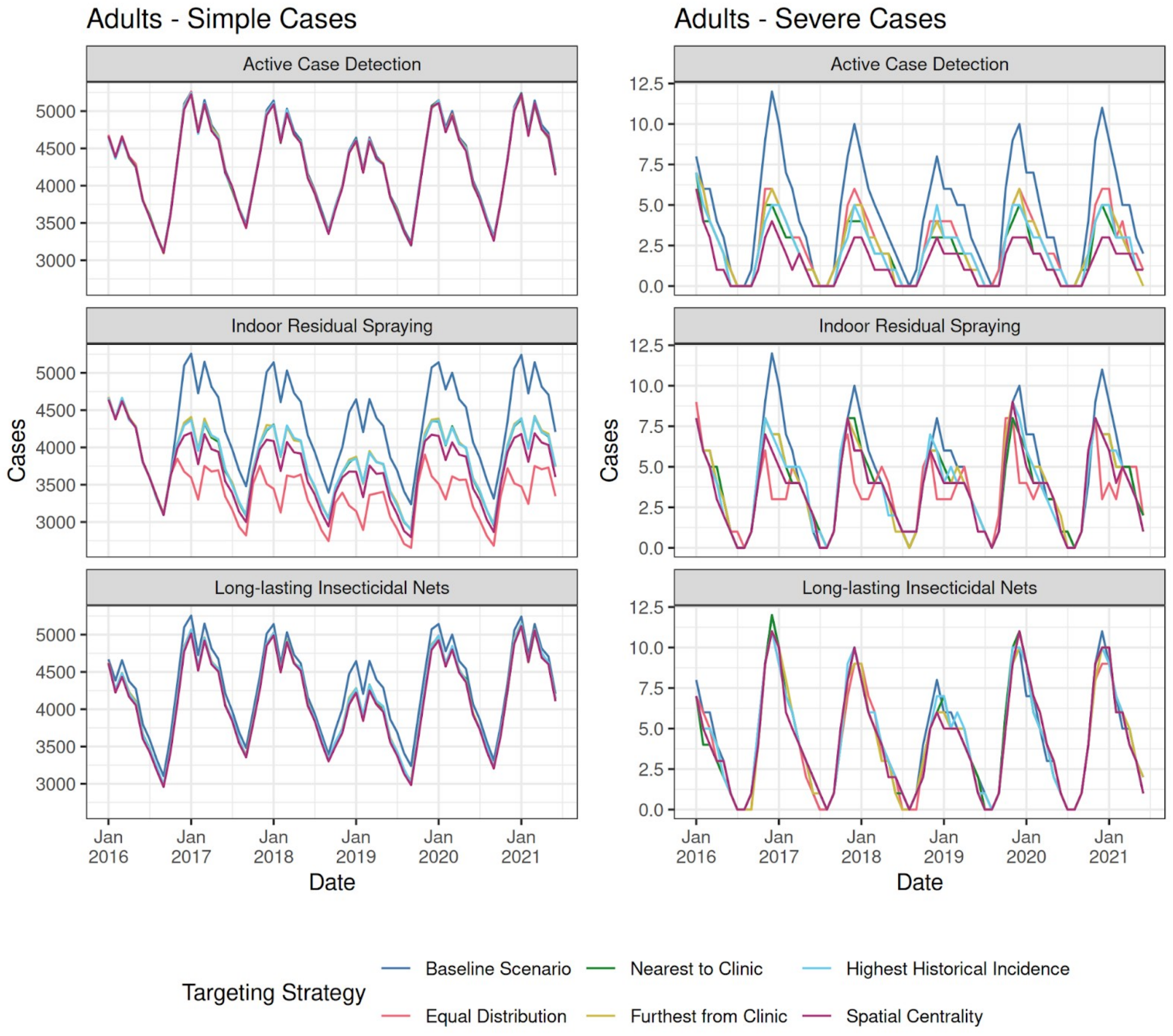


**Supplemental Figure 3.** Adult cases averted when applying one intervention by targeting strategy.

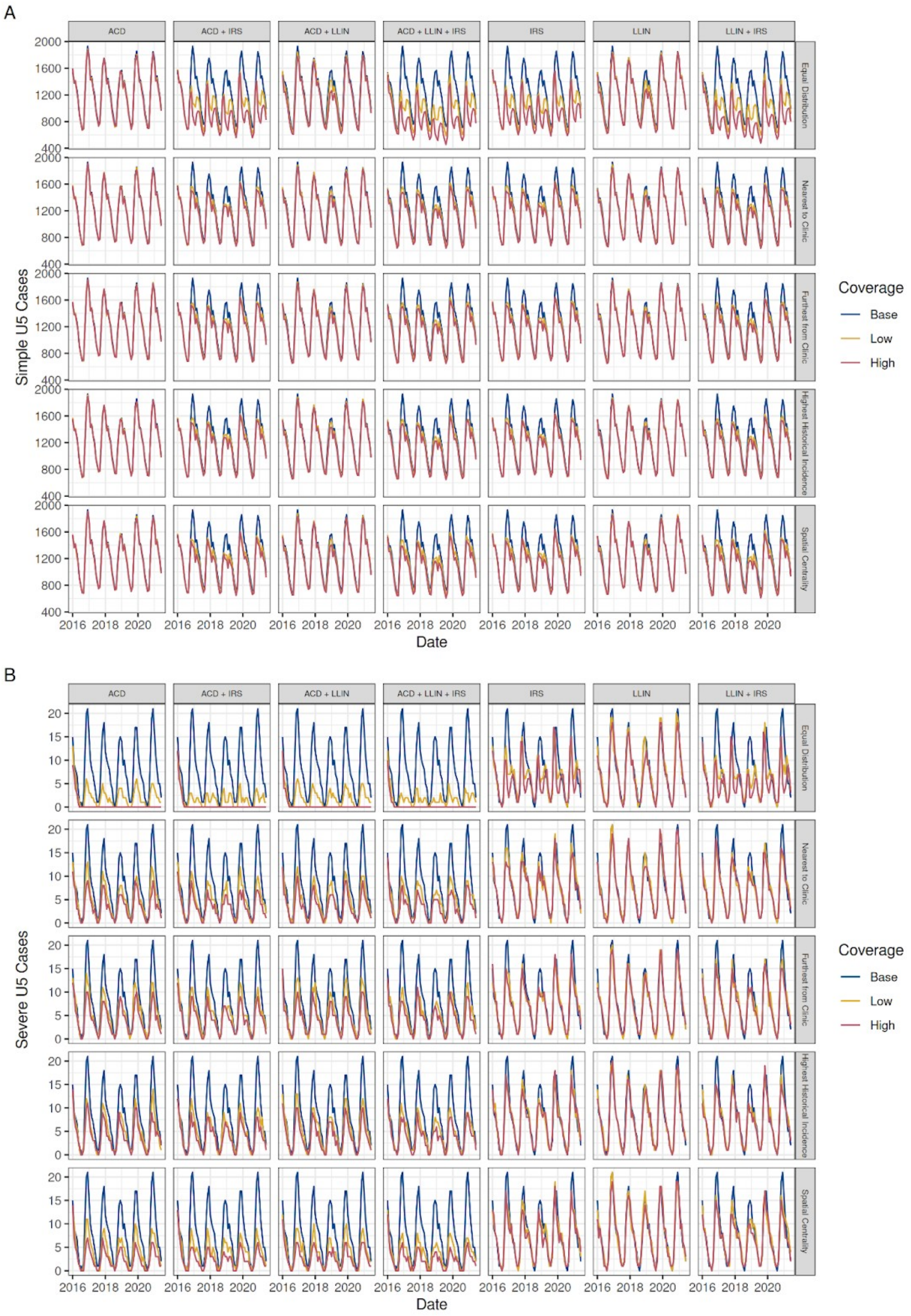


**Supplemental Figure 4.** Cases averted for children under 5 years old across differing combinations of interventions, targeting strategies, and coverage levels. Base coverage is the current coverage levels of interventions, as defined in the main manuscript. Low coverage is equal to 1.5x base levels or 100% coverage, whichever is lower. High coverage is equal to 2x base levels or 100% coverage, whichever is lower. Coverage is only increased in targeted fokontany, with other fokontany retaining base coverage levels.

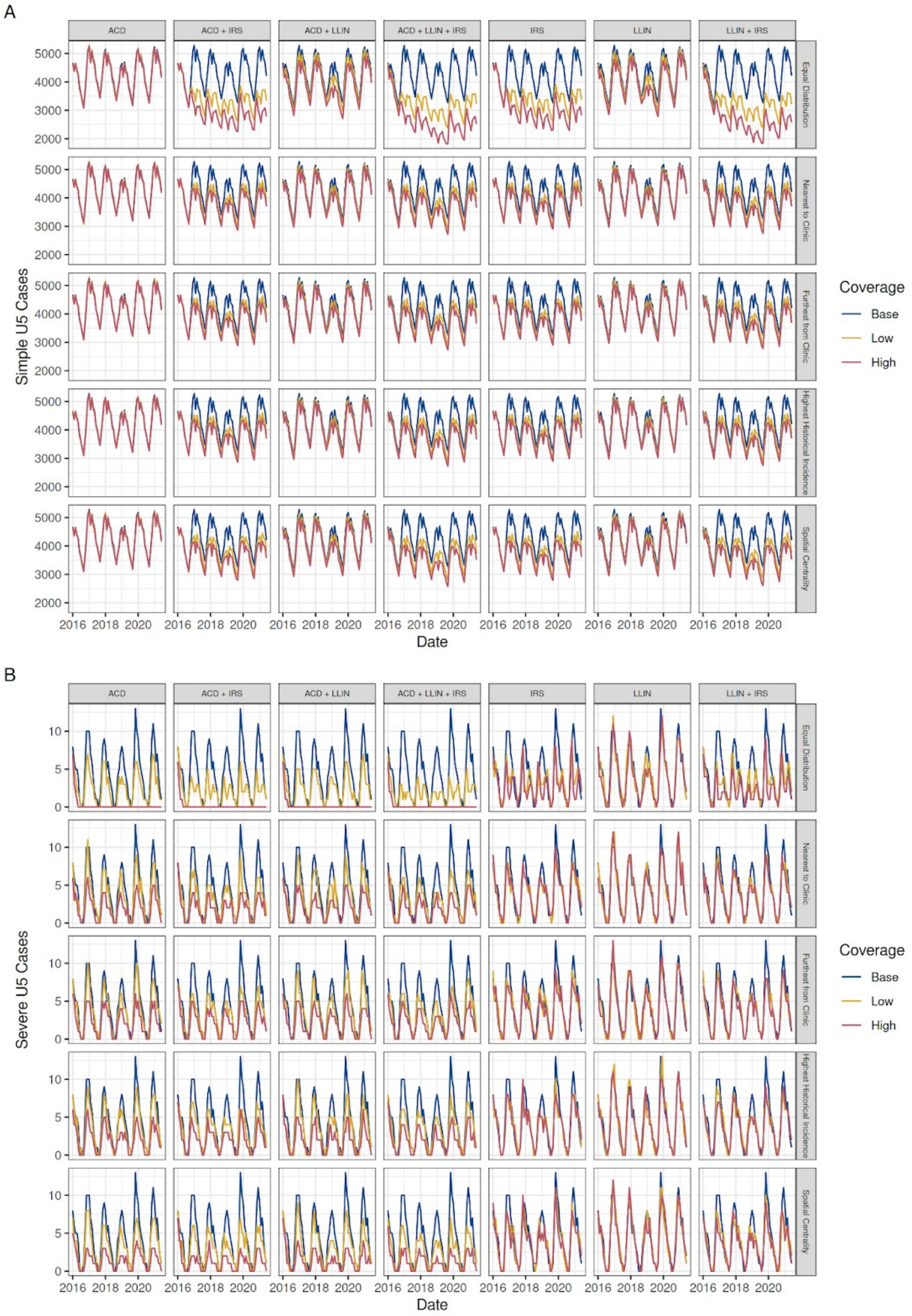


**Supplemental Figure 5.** Cases averted for adults across differing combinations of interventions, targeting strategies, and coverage levels. Base coverage is the current coverage levels of interventions, as defined in the main manuscript. Low coverage is equal to 1.5x base levels or 100% coverage, whichever is lower. High coverage is equal to 2x base levels or 100% coverage, whichever is lower. Coverage is only increased in targeted fokontany, with other fokontany retaining base coverage levels.
