## Supplemental Tables for "Leveraging spatial structure to design spatially-targeted malaria interventions at the community-scale"

**Supplemental Table 1. Transition rates between compartments and primary sources for values.**

| Symbol | Description | Value/Priors | Source |
|---|---|---|---|
| HOST EQUATION PARAMETERS | | | |
| $\gamma_A$ | Movement out of asymptomatic. Inverse of patent period of malaria | Prior: 0.0001-0.1 | Estimated |
| $\gamma_P$ | Movement from treated to susceptible. Inverse of protected period provided by prophylaxis | Prior: 0.0002-0.2 | Estimated |
| $\alpha$ | Movement from exposed to infected. Inverse of incubation period | 1/12 | Griffin et al. 2010 |
| $s^*$ | Age-dependent symptomatic rate | Children: 0.29<br>Adult: 0.089 | IHOPE 2021 |
| $\tau_s^*$ | Age-dependent treatment rate of symptomatic cases | Children: 0.57<br>Adult: 0.33 | IHOPE 2021 |
| $g^*$ | Age-dependent severe case rate | Children: 0.029<br>Adult: 0.006 | Kamau et al. 2020 |
| $\gamma_S$ | Movement from symptomatic to asymptomatic hosts. Inverse of clinical period. | 1/5 | Griffin et al. 2010 |
| $\tau_A^*$ | Age-dependent treatment rate of asymptomatic cases | Children: 0.0030<br>Adult: 0.0015 | Assuming 1 consultation per year for adults and 2 for children, with RDT sensitivity of 0.56. |
| $\gamma_G$ | Movement from severe to treated. Inverse of severe clinical period. | 1/14 | Bretscher et al. 2020 |
| $\beta_H^*$ | Age-dependent transmission rate from vector to host | Children: 0.59<br>Adult: 0.3 | Griffin et al. 2010 |

| Symbol | Description | Value/Priors | Source |
|---|---|---|---|
| VECTOR EQUATION PARAMETERS | | | |
| $b(T)$ | Eggs per day | f(Temperature) | Mordecai et al. 2013 |
| $\varepsilon(T)$ | Larval development rate | f(Temperature) | Mordecai et al. 2013 |
| $\mu_A(T)$ | Adult mortality rate | f(Temperature) | Mordecai et al. 2013 |
| $\Delta(T)$ | Parasite development rate | f(Temperature) | Mordecai et al. 2013 |
| $a(T)$ | Bite rate | f(Temperature) | Mordecai et al. 2013 |
| $\beta_V$ | Class-dependent transmission rate from host to vector. | Prior:<br>Symptomatic:<br>0.001 – 0.3 | Estimated symptomatic rate.<br>Asymptomatic: Held at 30% of symptomatic<br>Treated: Held at 5% of symptomatic |
| $L_f$ | Fractional larval growth state | 0.5 | Approximates average larval life stage |
| $K_{flush}$ | Maximum mortality rate due to flushing from rainfall | 0.4 | Tompkins and Ermert 2013 |
| $T_{flush}$ | Scaling parameter to define the shape of the relationship between rainfall and larval mortality | 0.5 | Tompkins and Ermert 2013 |
| $K$ | Carrying capacity of flooded rice field (individuals/m$^2$) | Prior:<br>10 - 35 | Estimated |

**Supplemental Table 1 References**


Bretscher, M. T., P. Dahal, J. Griffin, K. Stepniewska, Q. Bassat, E. Baudin, U. D’Alessandro, A. A. Djimde, G. Dorsey, E. Espié, B. Fofana, R. González, E. Juma, C. Karema, E. Lasry, B. Lell, N. Lima, C. Menéndez, G. Mombo-Ngoma, C. Moreira, F. Nikiema, J. B. Ouédraogo, S. G. Staedke, H. Tinto, I. Valea, A. Yeka, A. C. Ghani, P. J. Guerin, and L. C. Okell. 2020. The duration of chemoprophylaxis against malaria after treatment with artesunate-amodiaquine and artemether-lumefantrine and the effects of pfmdr1 86Y and pfcrt 76T: a meta-analysis of individual patient data. BMC Medicine 18:47.

Griffin, J. T., T. D. Hollingsworth, L. C. Okell, T. S. Churcher, M. White, W. Hinsley, T. Bousema, C. J. Drakeley, N. M. Ferguson, M.-G. Basáñez, and A. C. Ghani. 2010. Reducing Plasmodium falciparum Malaria Transmission in Africa: A Model-Based Evaluation of Intervention Strategies. PLOS Medicine 7:e1000324.

Kamau, A., G. Mtanje, C. Mataza, G. Mwambingu, N. Mturi, S. Mohammed, G. Ong’ayo, G. Nyutu, A. Nyaguara, P. Bejon, and R. W. Snow. 2020. Malaria infection, disease and mortality among children and adults on the coast of Kenya. Malaria Journal 19.

Mordecai, E. A., K. P. Paaijmans, L. R. Johnson, C. Balzer, T. Ben-Horin, E. de Moor, A. McNally, S. Pawar, S. J. Ryan, T. C. Smith, and K. D. Lafferty. 2013. Optimal temperature for malaria transmission is dramatically lower than previously predicted. Ecology Letters 16:22–30.

Tompkins, A. M., and V. Ermert. 2013. A regional-scale, high resolution dynamical malaria model that accounts for population density, climate and surface hydrology. Malaria Journal 12:65.

**Supplemental Table 2. Variation in Transition Rates due to Environmental Forcing**

| Environmental Variable | Parameter | Minimum Daily Variation | Median Daily Variation | Maximum Daily Variation | Mean Daily Coefficient of Variation |
|---|---|---|---|---|---|
| Flooding | Mosquito Carrying Capacity (millions), W**K* | 4.5998 | 9.6621 | 14.9341 | 0.8328 |
| Temperature | Parasite Development Rate , $\Delta(T)$ | 0.0382 | 0.0485 | 0.0548 | 0.1559 |
| Temperature | Larval Development Rate, $\varepsilon(T)$ | 0.0364 | 0.0472 | 0.0537 | 0.1545 |
| Temperature | Eggs per Female per Day, $b(T)$ | 2.7743 | 5.2403 | 8.6745 | 0.1431 |
| Temperature | Bite rate, $a(T)$ | 0.1145 | 0.1345 | 0.1656 | 0.1198 |
| Temperature | Larval Mosquito Survival, $\mu_L(T)$ | 0.0618 | 0.2592 | 0.6927 | 0.0993 |
| Rainfall | Flushing-Induced Larval Mortality, $\mu_L(R)$ | 0 | 0 | 0.2989 | 0.0063 |
| Temperature | Adult Mosquito Survival, $\mu_A(T)$ | 0.0056 | 0.0174 | 0.0528 | 0.0041 |

**Supplemental Table 3. Averted cases for combined interventions at high and low coverage levels.**

| | | Interventions | | | | Targeting Scenario | | | | |
|---|---|---|---|---|---|---|---|---|---|---|
| | | IRS | LLIN | ACS | Coverage Level | Equally Distributed | Nearest CSBs | Furthest from CSBs | Historical Incidence | Spatial Centrality |
| Children Under 5 | Simple Cases | X | | | low | 2559 (2411 - 2786) | 942 (897 - 989) | 908 (860 - 944) | 950 (913 - 968) | 1194 (1107 - 1282) |
| | | X | | | high | 3982 (3821 - 4408) | 1327 (1232 - 1455) | 1290 (1210 - 1418) | 1293 (1192 - 1432) | 1666 (1522 - 1833) |
| | | | X | | low | 382 (150 - 660) | 203 (83 - 329) | 176 (29 - 346) | 185 (63 - 319) | 224 (56 - 461) |
| | | | X | | high | 848 (326 - 1558) | 376 (159 - 627) | 365 (178 - 591) | 365 (162 - 628) | 441 (168 - 832) |
| | | | | X | low | 279 (255 - 327) | 129 (81 - 184) | 132 (106 - 160) | 155 (111 - 191) | 183 (104 - 250) |
| | | | | X | high | 354 (287 - 398) | 215 (139 - 260) | 164 (103 - 224) | 201 (140 - 243) | 242 (201 - 306) |
| | | X | X | | low | 3153 (2895 - 3379) | 1117 (1014 - 1271) | 1080 (992 - 1172) | 1083 (950 - 1216) | 1351 (1270 - 1458) |
| | | X | X | | high | 5260 (4765 - 5618) | 1622 (1435 - 1770) | 1540 (1312 - 1704) | 1581 (1342 - 1782) | 2032 (1818 - 2232) |
| | | | X | X | low | 659 (461 - 910) | 320 (161 - 479) | 296 (178 - 465) | 294 (194 - 424) | 380 (229 - 595) |
| | | | X | X | high | 1172 (664 - 1811) | 530 (303 - 820) | 538 (335 - 767) | 538 (326 - 825) | 676 (406 - 1008) |

| | | | | | | | | | | |
|---|---|---|---|---|---|---|---|---|---|---|
| | | X | | X | low | 2786 (2675 - 2991) | 1116 (1005 - 1232) | 1042 (997 - 1078) | 1048 (1000 - 1096) | 1343 (1268 - 1393) |
| | | X | | X | high | 4254 (4070 - 4638) | 1509 (1441 - 1568) | 1444 (1336 - 1569) | 1461 (1389 - 1557) | 1891 (1799 - 2070) |
| | | X | X | X | low | 3364 (3081 - 3596) | 1244 (1092 - 1396) | 1192 (1071 - 1334) | 1209 (1117 - 1339) | 1527 (1376 - 1700) |
| | | X | X | X | high | 5511 (5074 - 5884) | 1778 (1601 - 1975) | 1710 (1528 - 1864) | 1722 (1522 - 1864) | 2270 (2014 - 2464) |
| | Severe Cases | X | | | low | 17 (14 - 19) | 5 (3 - 7) | 8 (5 - 12) | 7 (4 - 10) | 8 (3 - 12) |
| | | X | | | high | 27 (24 - 30) | 8 (7 - 10) | 10 (6 - 14) | 8 (3 - 12) | 13 (11 - 16) |
| | | | X | | low | 0 (0 - 3) | 0 (0 - 4) | 5 (2 - 8) | 3 (0 - 5) | 0 (0 - 2) |
| | | | X | | high | 8 (4 - 16) | 2 (0 - 6) | 5 (1 - 7) | 4 (0 - 8) | 2 (0 - 5) |
| | | | | X | low | 71 (67 - 74) | 39 (37 - 41) | 33 (32 - 35) | 37 (33 - 44) | 48 (45 - 50) |
| | | | | X | high | 95 (87 - 100) | 52 (51 - 54) | 47 (43 - 50) | 48 (43 - 51) | 66 (59 - 72) |
| | | X | X | | low | 22 (20 - 23) | 10 (8 - 10) | 8 (4 - 11) | 8 (4 - 11) | 10 (4 - 13) |
| | | X | X | | high | 34 (31 - 37) | 10 (9 - 10) | 11 (8 - 12) | 10 (10 - 12) | 15 (12 - 17) |
| | | | X | X | low | 71 (67 - 76) | 42 (37 - 45) | 31 (29 - 34) | 35 (31 - 39) | 49 (46 - 52) |
| | | | X | X | high | 95 (87 - 100) | 53 (51 - 54) | 45 (43 - 46) | 49 (46 - 52) | 64 (58 - 67) |
| | | X | | X | low | 77 (68 - 83) | 43 (36 - 49) | 36 (32 - 41) | 41 (34 - 46) | 51 (49 - 54) |
| | | X | | X | high | 95 (87 - 100) | 56 (51 - 62) | 48 (43 - 52) | 50 (45 - 53) | 68 (63 - 71) |
| | | X | X | X | low | 77 (73 - 80) | 42 (42 - 43) | 36 (33 - 39) | 40 (37 - 45) | 53 (48 - 55) |

| | | | | | | | | | | |
|---|---|---|---|---|---|---|---|---|---|---|
| | | X | X | X | high | 95 (87 - 100) | 57 (53 - 61) | 47 (45 - 50) | 52 (50 - 54) | 70 (65 - 75) |
| Adults | Simple Cases | X | | | low | 12063 (11514 - 12378) | 4952 (4749 - 5259) | 4784 (4649 - 5024) | 4871 (4673 - 5174) | 5999 (5761 - 6351) |
| | | X | | | high | 17410 (16826 - 17853) | 6580 (6368 - 6878) | 6390 (6219 - 6684) | 6539 (6311 - 6807) | 8123 (7886 - 8531) |
| | | | X | | low | 2157 (1534 - 3570) | 1068 (714 - 1774) | 983 (678 - 1706) | 1022 (610 - 1773) | 1232 (837 - 2130) |
| | | | X | | high | 4487 (3415 - 7214) | 2048 (1437 - 3294) | 1964 (1499 - 3129) | 2022 (1563 - 3259) | 2517 (1904 - 4058) |
| | | | | X | low | 336 (235 - 502) | 194 (163 - 248) | 130 (18 - 270) | 143 (28 - 231) | 186 (93 - 251) |
| | | | | X | high | 763 (637 - 820) | 456 (399 - 499) | 355 (292 - 475) | 367 (237 - 504) | 524 (470 - 589) |
| | | X | X | | low | 14418 (14171 - 14774) | 5666 (5369 - 5906) | 5503 (5224 - 5747) | 5662 (5445 - 5902) | 6954 (6695 - 7212) |
| | | X | X | | high | 22065 (20792 - 23152) | 7896 (7474 - 8285) | 7698 (7295 - 8027) | 7768 (7472 - 8086) | 9832 (9419 - 10369) |
| | | | X | X | low | 2412 (1866 - 3869) | 1151 (795 - 1881) | 1180 (820 - 1899) | 1186 (860 - 1864) | 1434 (1080 - 2297) |
| | | | X | X | high | 5200 (4227 - 7858) | 2448 (2014 - 3671) | 2374 (1862 - 3587) | 2363 (1834 - 3572) | 2916 (2306 - 4409) |
| | | X | | X | low | 12233 (11631 - 12494) | 5052 (4847 - 5342) | 4926 (4748 - 5258) | 4986 (4757 - 5337) | 6182 (5918 - 6466) |
| | | X | | X | high | 17870 (17293 - 18315) | 6938 (6717 - 7311) | 6687 (6500 - 6941) | 6852 (6729 - 7097) | 8594 (8335 - 8967) |
| | | X | X | X | low | 14683 (14418 - | 5826 (5515 - | 5660 (5486 - | 5790 (5597 - 5972) | 7136 (6909 - |

| | | | | | | | | | | |
|---|---|---|---|---|---|---|---|---|---|---|
| | | | | | | 15126) | 6104) | 5772) | | 7377) |
| | | X | X | X | high | 22572 (21278 - 23603) | 8241 (7921 - 8632) | 7922 (7640 - 8299) | 8074 (7800 - 8408) | 10288 (9957 - 10757) |
| | Severe Cases | X | | | low | 14 (11 - 17) | 7 (2 - 12) | 6 (4 - 10) | 7 (3 - 11) | 5 (1 - 10) |
| | | X | | | high | 19 (16 - 24) | 6 (3 - 8) | 10 (7 - 15) | 8 (2 - 15) | 7 (2 - 12) |
| | | | X | | low | 2 (0 - 8) | 1 (0 - 8) | 3 (1 - 7) | 1 (0 - 5) | 2 (0 - 6) |
| | | | X | | high | 4 (0 - 10) | 1 (0 - 7) | 1 (0 - 8) | 1 (0 - 8) | 2 (0 - 9) |
| | | | | X | low | 23 (20 - 27) | 13 (9 - 17) | 11 (9 - 13) | 11 (6 - 16) | 16 (13 - 21) |
| | | | | X | high | 53 (45 - 59) | 31 (24 - 38) | 26 (19 - 31) | 29 (20 - 35) | 36 (31 - 42) |
| | | X | X | | low | 16 (10 - 21) | 7 (3 - 11) | 6 (0 - 12) | 7 (2 - 13) | 8 (3 - 13) |
| | | X | X | | high | 24 (22 - 27) | 9 (4 - 15) | 9 (5 - 14) | 10 (6 - 16) | 10 (7 - 15) |
| | | | X | X | low | 23 (19 - 27) | 13 (8 - 19) | 10 (7 - 18) | 13 (9 - 19) | 16 (14 - 21) |
| | | | X | X | high | 53 (45 - 59) | 30 (26 - 32) | 28 (20 - 32) | 30 (24 - 37) | 38 (33 - 44) |
| | | X | | X | low | 29 (25 - 34) | 16 (14 - 19) | 15 (9 - 20) | 16 (11 - 21) | 18 (13 - 24) |
| | | X | | X | high | 53 (45 - 59) | 32 (28 - 37) | 27 (20 - 33) | 30 (23 - 33) | 40 (33 - 47) |
| | | X | X | X | low | 32 (26 - 40) | 18 (15 - 21) | 15 (11 - 21) | 15 (12 - 20) | 22 (17 - 26) |
| | | X | X | X | high | 53 (45 - 59) | 32 (27 - 35) | 28 (26 - 31) | 31 (27 - 36) | 39 (33 - 46) |
